# Comparative efficacy and safety of pharmacological interventions for the treatment of COVID-19: A systematic review and network meta-analysis of confounder-adjusted 20212 hospitalized patients

**DOI:** 10.1101/2020.06.15.20132407

**Authors:** Min Seo Kim, Min Ho An, Won Jun Kim, Tae-Ho Hwang

## Abstract

**Objective:** To evaluate the comparative efficacy and safety of pharmacological interventions used in treating COVID-19 and form a basis for an evidence-based guideline of COVID-19 management by evaluating the level of evidence behind each treatment regimen in different clinical settings.

**Design:** Systematic review and network meta-analysis

**Data Sources:** PubMed, Google Scholar, MEDLINE, the Cochrane Library, medRxiv, SSRN, WHO International Clinical Trials Registry Platform, and ClinicalTrials.gov up to June 9^th^, 2020.

**Study Selection:** Published and unpublished randomized controlled trials (RCTs) and baseline-adjusted observational studies which met our predefined eligibility criteria.

**Main Outcome Measures:** The outcomes of interest were mortality, progression to severe disease (severe pneumonia or admission to intensive care unit (ICU)), time to viral clearance, QT prolongation, fatal cardiac complications, and non-cardiac serious adverse events. The level of evidence behind each outcome was also measured using the Grading of Recommendations Assessment, Development, and Evaluation (GRADE) framework.

**Results:** 49 studies with a total of 20212 confounder-adjusted patients were included for analysis. The risk of progression to severe pneumonia or ICU admission was significantly reduced with tocilizumab (GRADE low), anakinra (GRADE very low), and remdesivir (GRADE high) compared to standard care. Tocilizumab was shown to reduce mortality rate for both moderate-severe patients in the non-ICU setting at admission (Odds ratio (OR) 0.31, 95% confidence interval (CI) 0.18 to 0.54, GRADE low) and critically ill patients in the ICU setting (OR 0.67, 95% CI 0.50 to 0.91, GRADE low). High dose IVIG reduced death rate (GRADE low) while corticosteroids increased mortality for critically ill patients (GRADE moderate). Convalescent plasma and hydroxychloroquine were shown to promote viral clearance (OR 11.39, 95% CI 3.91 to 33.18, GRADE low and OR 6.08, 95% CI 2.74 to 13.48, GRADE moderate, respectively) while not altering mortality or progression to the severe courses. The combination of hydroxychloroquine and azithromycin was shown to be associated with increased QT prolongation incidence (OR 1,85, 95% CI 1.05 to 3.26, GRADE low) and fatal cardiac complications in cardiac-impaired populations (OR 2.26, 95% CI 1.26 to 4.05, GRADE low). High-dose (>600mg/day) hydroxychloroquine monotherapy was significantly associated with increased non-cardiac serious adverse events (GRADE moderate).

**Conclusion:** Anti-inflammatory agents (tocilizumab, anakinra, and IVIG) and remdesivir may safely and effectively improve outcomes of hospitalized COVID-19 patients. Widely used hydroxychloroquine provides marginal clinical benefit in improving viral clearance rates whilst posing both cardiac and non-cardiac safety risks, especially in the vulnerable population. Only 20% of current evidence on pharmacological management of COVID-19 is on moderate and high evidence certainty and can be considered in practice and policy; remaining 80% are of low or very low certainty and warrant further studies to establish firm conclusions.

**Systematic Review Registration:** PROSPERO 2020: CRD42020186527.

**Summary Box:** *Section 1: What is already known on this topic:*

- Numerous clinical trials and observational studies have investigated various pharmacological agents as potential treatment for COVID-19.
- Results from numerous studies are heterogeneous and sometimes even contradictory to one another, making it difficult for clinicians to determine which treatments are truly effective.
- Level of evidence behind each outcome from diverse studies remains unknown.

*Section 2: What this study adds:*

- Anti-inflammatory agents (tocilizumab, anakinra, and IVIG) and remdesivir may safely and effectively improve clinical outcomes of COVID-19.
- Widely used hydroxychloroquine provides marginal clinical benefit in improving viral clearance rates whilst posing both cardiac and non-cardiac safety risks.
- Only 20% of current evidence on pharmacological management of COVID-19 is on moderate/high evidence certainty and can be considered in practice and policy; remaining 80% are of low or very low certainty and warrant further studies to establish firm conclusions.

## Introduction

A large registry-based study investigating the effect of hydroxychloroquine on COVID-19 patients reported surprisingly high mortality rate and ventricular arrhythmia incidence with hydroxychloroquine use^1^, and this paper contributed to the decision of the World Health Organization (WHO) to pause all ongoing trials on hydroxychloroquine due to safety concerns. Although this paper was retracted after concerns raised by clinicians and scientists, the delay of trials and confusion caused by the paper were unavoidable. This was an alarming event that due to the sudden advent of COVID-19 and global urgency to find treatments, many clinical and observational studies are of suboptimal quality^2^; therefore, it may be unreliable to make decisions grounded on the evidence of a single paper. Prospective meta-analyses synthesizing multiple studies using predefined eligibility criteria can be an interim solution to generate reliable conclusions^2^ and protect from ill-informed changes in practice and policy.

Numerous COVID-19 clinical trials are underway, and over 31 pharmacological agents and combinations have been investigated as potential treatments of COVID-19 to date. Network meta-analysis (NMA) is an analytical tool that enables the a single coherent ranking of such numerous interventions, and it can thus aid decision-makers who must choose amongst an array of treatment options ^3^.

We conducted the first network meta-analysis with selective predefined eligibility criteria for both published and unpublished data, and investigated 31 treatment regimens for comparative efficacy and safety. We incorporated 49 studies (16 randomized controlled trials (RCT) and 33 baseline-adjusted observational studies) including a total of 20212 confounder-adjusted patients. The level of certainty behind the evidence for each outcome was evaluated to assist the decision-making of clinicians and policy makers. This study will serve as the basis for an individual patient data (IPD) network meta-analysis that we are designing as a future study.

## Methods

### Search strategy and selection criteria

We searched PubMed, Google Scholar, MEDLINE, the Cochrane Library, medRxiv, SSRN, WHO International Clinical Trials Registry Platform, and ClinicalTrials.gov for RCTs and observational studies that evaluated treatment responses to pharmacological management in COVID-19 patients, from inception to June 9^th^, 2020. Reference lists of review articles were also reviewed to search for additional articles that may not have been retrieved by the prespecified searching strategy. We had no restriction on language, but all included studies were written in English.

We contacted principal investigators of unpublished studies identified in trial registries and regulatory submissions to obtain unpublished data. Inclusion of unpublished data in NMAs is not uncommon ^4-11^ and reduces risk of selection and publication bias while increasing the density of study data. This is especially beneficial in the study of COVID-19 as all data were generated relatively recently, and the bulk of the relevant data are still in the unpublished stages. Pre-prints have been used in meta-analysis relatively frequently for the urgent topic of COVID-19^9-12^, and American Gastroenterological Association (AGA) recently published a management guideline for the gastrointestinal manifestation of COVID-19 patients based on the result of meta-analysis incorporating pre-prints^11^. We contacted authors of included pre-prints from medRxiv and SSRN, and any change in the results was updated.

We included both RCTs and baseline-adjusted observational studies; the rationale is that inclusion of real-world data from non-randomized studies has the potential to improve precision of findings from RCTs if appropriately integrated^13^ and that the volume of information provided by these studies is necessary to assess adverse events of low to moderate incidence^14-16^. As observational studies are more vulnerable to bias, we included only the studies that adjusted for relevant confounding variables through methods such as propensity score matching (PSM), inverse probability treatment weighting (IPTW), or regression model adjustment. Studies providing evidence that the risk for such confounding was low by establishing baseline similarity between the groups also met inclusion criteria (Appendix p5).

Following studies were excluded: studies without a proper control group; studies of children or adolescents (<18 years); observational studies with significant differences in baseline characteristics between groups and did not perform adequate adjustments; studies investigating the effect of medication initiated prior to the diagnosis of COVID-19 (e.g. ACEi/ARB for hypertensive patients).

### Data extraction and quality assessment

The study search and data extraction were independently conducted by 3 authors (MS Kim, MH An, and WJ Kim). Manuscript and supplementary materials of the included studies were reviewed for relevant information which was extracted according to a pre-specified protocol. Any discrepancy or ambiguity in this process was resolved by discussion. Authors of certain included studies were contacted in case of missing or unclear information. Non-randomized studies were qualitatively assessed using the Newcastle-Ottawa Scale (NOS)^17^, and RCTs were assessed with the Jadad scale^18^. All studies were assessed for risk of bias using the RoB 2 tool for randomized studies and ROBINS-I tool for nonrandomized studies^19^. The quality of evidence of collective outcomes were estimated using the Grading of Recommendations Assessment, Development, and Evaluation (GRADE) framework^20^. A comparison-adjusted funnel plot with Egger’s test was constructed to assess for publication bias^21^.

Control groups consisted of patients who received standard care or placebo. Patients who received hydroxychloroquine or corticosteroids were subdivided according to the dosage they received. For hydroxychloroquine, most studies reported 400mg hydroxychloroquine daily for maintenance, and this was considered the standard prescription; patients who received daily maintenance dosage of over 600mg hydroxychloroquine were classified into a separate high-dose hydroxychloroquine group. For corticosteroids, average daily dosage of 40mg methylprednisolone (or equivalent) was regarded as the standard dosage, while 1-2mg/kg/day methylprednisolone (or equivalent) was regard as high dose. 1mg methylprednisolone was considered equivalent to 0.1875mg dexamethasone and 5mg hydrocortisone.

A critically ill patient was defined as a patient who received invasive mechanical ventilation or needed intensive care in the ICU before or soon after beginning the treatment of interest, while moderate-severe patients were defined as patients hospitalized in a non-ICU setting at admission. The mortality rate of patients included in our mortality analyses were 11.7% for moderate-severe (non-ICU) patients and 38.6% for critically ill (ICU) patients on average.

### Data synthesis and statistical analysis

We conducted a random-effects network meta-analysis within a frequentist framework using STATA (Stata Corp, College Station, TX, US, version 15.0) and R (version 3.6.0) software^22^. Direct and indirect (and mixed) comparison were accomplished through the self-programmed routines of STATA^21 23^ and the netmeta package of R^24^, as done in our previous work^25^. The effect estimation was in odds ratios (OR) for dichotomous variables and mean difference (MD) for continuous variables, both with 95% confidential intervals (CI). When median (interquartile range) was presented for continuous variables of interest, it was converted to mean (standard deviation) by calculation^26 27^. A two-sided p-value of less than 0.05 was regarded as statistically significant.

Statistical heterogeneity was estimated using restricted maximum likelihood method^28^ and expressed with Higgins I^2^ statistics and the Cochran Q test^29^. The net heat plot was constructed to visualize the inconsistency matrix and detect specific comparisons which introduced large inconsistencies^30^. The rank of effect estimation for each treatment was investigated using the surface under the cumulative rank curve (SUCRA) of P rank score of R31.

Prespecified subgroup and sensitivity analyses were performed to determine whether the results were affected by the patient severity, treatment protocol, and study design. The primary outcomes were separately analyzed for moderate to severe patients (non-ICU at admission) and critically ill patients (ICU) as these patients may respond differently to treatments. Sensitivity analyses were conducted by restricting the analyses to only RCTs, only published studies, excluding studies with high/serious risk of bias, and excluding studies in which initiation of treatment was over 14 days after symptom onset.

### Patient and public involvement

Neither any patients nor the public were involved in the design, conduct, and reporting of the research. The study protocol is publicly available on PROSPERO (CRD42020186527) and medRxiv.

## Results

The initial search identified 5970 articles. These studies were assessed for inclusion using the prespecified inclusion and exclusion criteria described in methods. Title and abstract of 3,626 articles were assessed, and 251 studies were found suitable for full-text review. After excluding 202 studies, 16 RCTs and 33 baseline-adjusted observations studies were finally included in our network meta-analysis (Figure 1). Total of 20212 confounder-adjusted COVID-19 patients were included. Background characteristics and reference list of included studies are presented in the supplementary appendix pp113-170. The risk of bias in included studies were generally low to moderate (Supplementary appendix pp 63-108).

**Figure 1.**
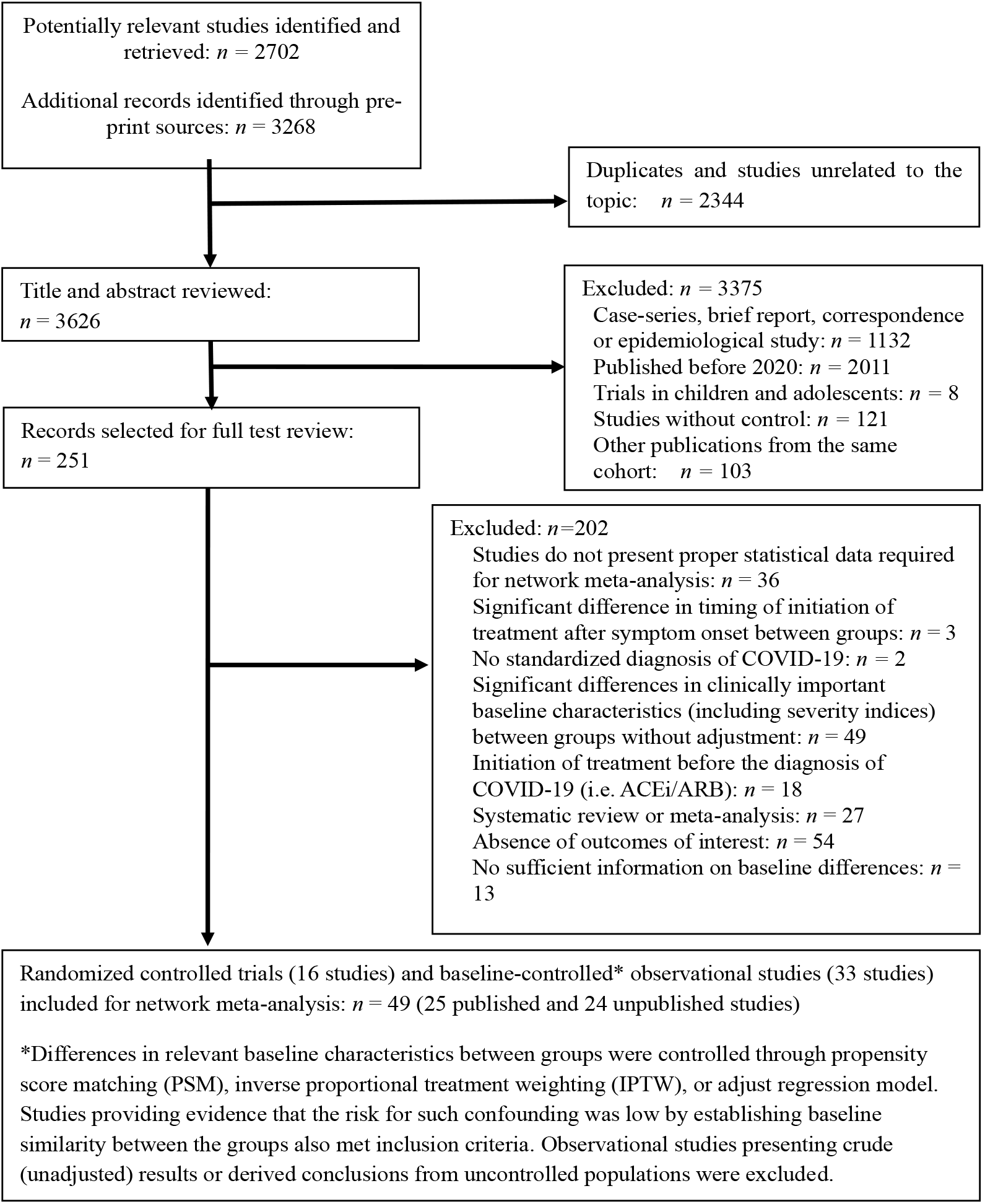
PRISMA diagram showing selection of articles for pairwise and network meta-analysis

For both pairwise meta-analysis and network meta-analysis, the primary outcomes presented no evidence of heterogeneity (Appendix pp 12-47). Inconsistency, which represents discordance of direct and indirect comparisons, was also evaluated for outcomes, but none were subject to global inconsistency. The network of eligible comparisons for clinically relevant outcomes are presented in Figure 2. Detailed information of studies included in the analysis for cardiac adverse events are presented in Table 1, and the certainty of evidence (GRADE) for each outcome is summarized in Table 2.

**Table 1.**
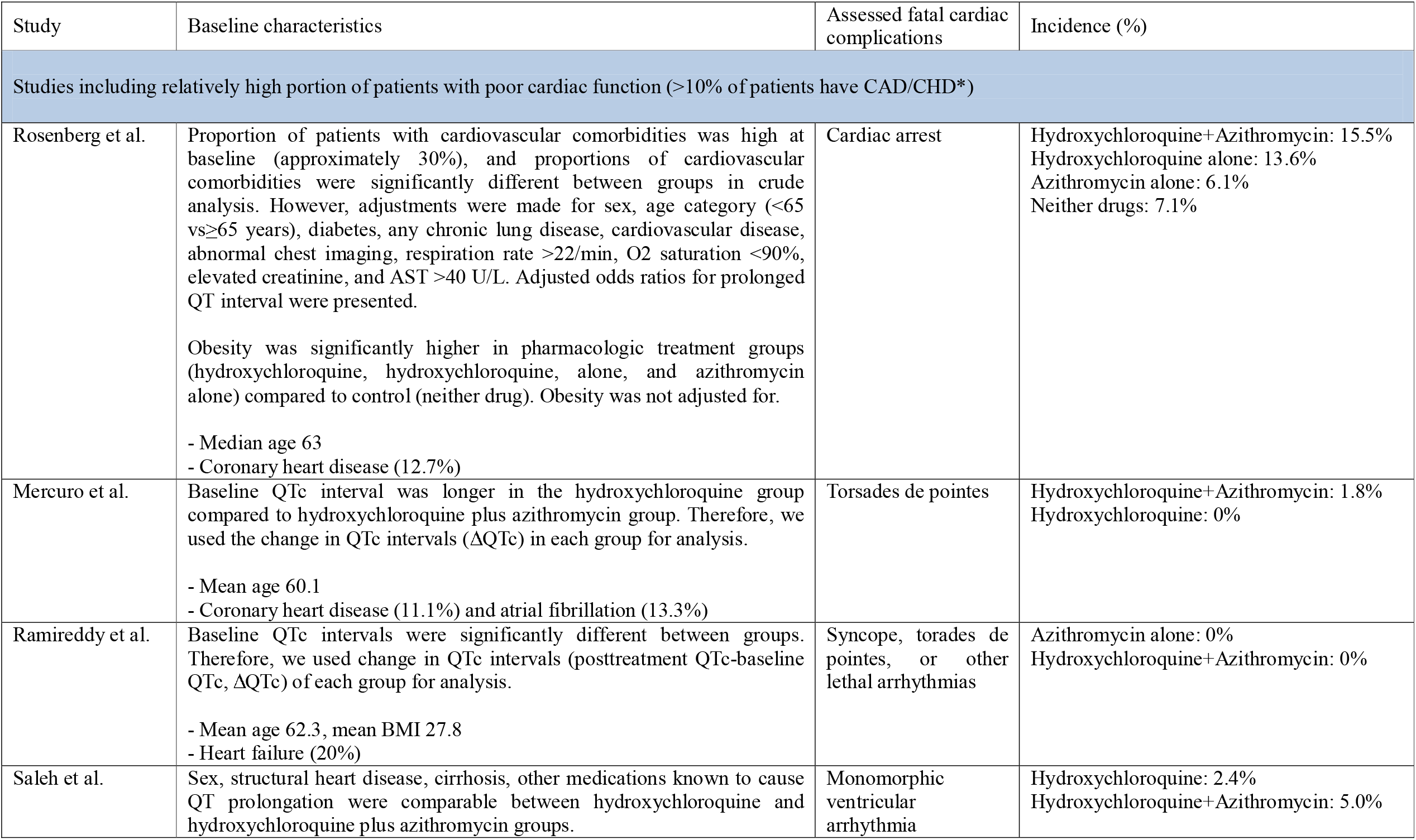
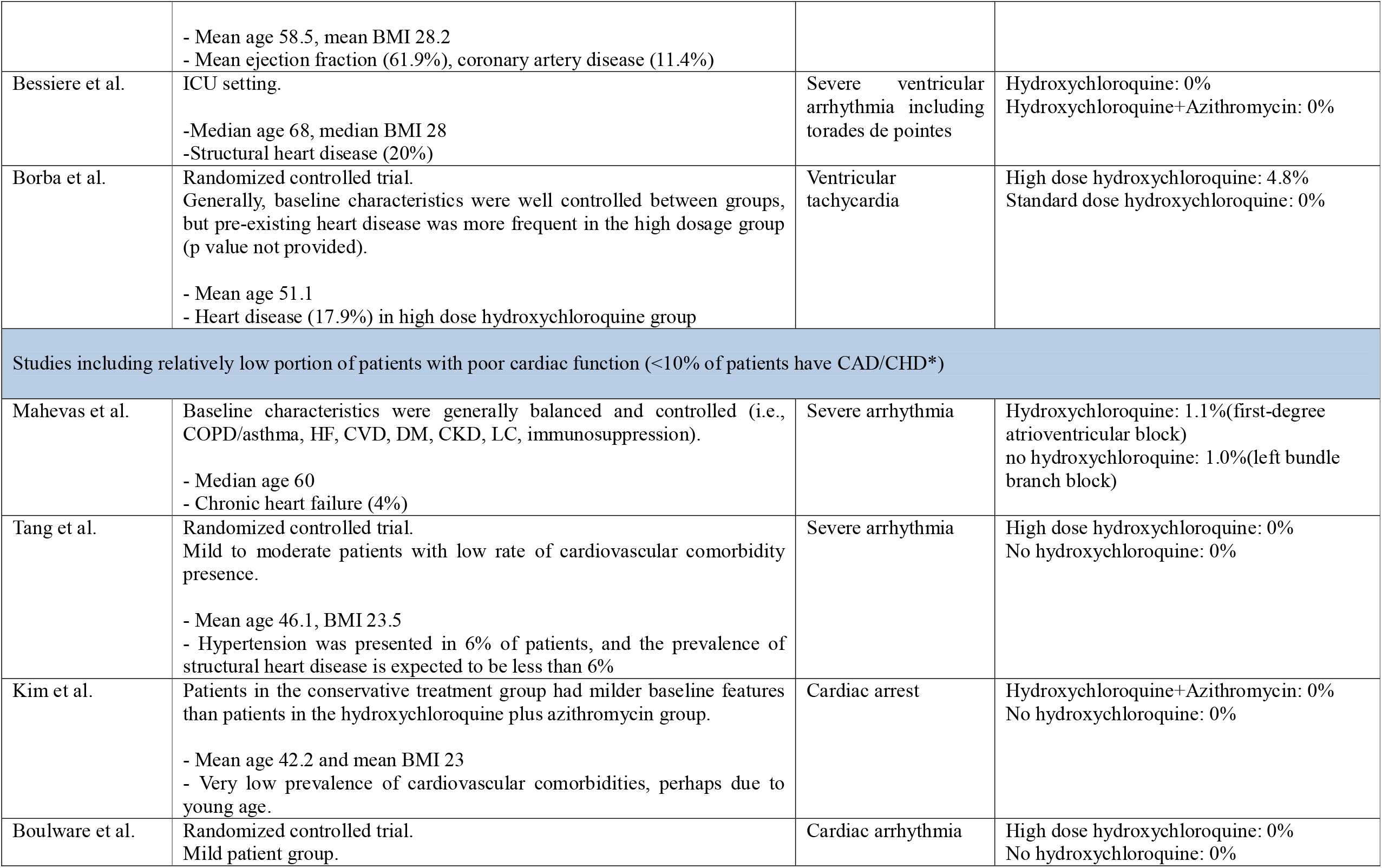
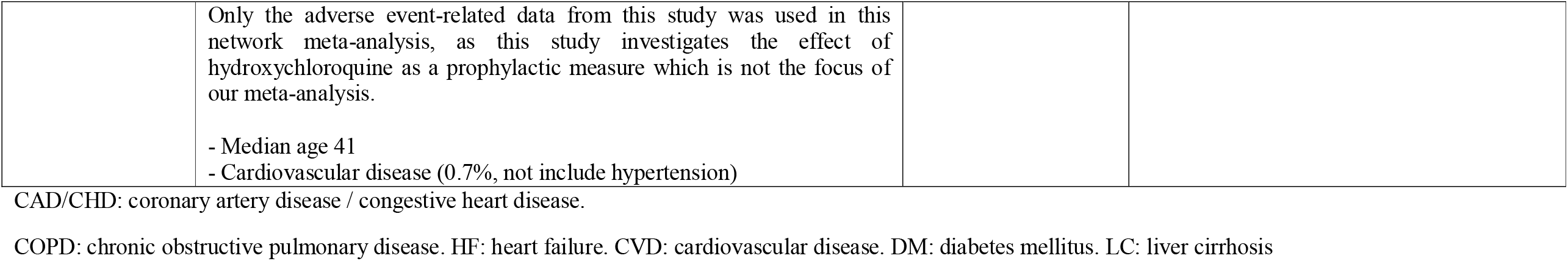
Studies included for analysis of QT prolongation and fatal cardiac complications after taking hydroxychloroquine alone or hydroxychloroquine with azithromycin.

**Table 2.**
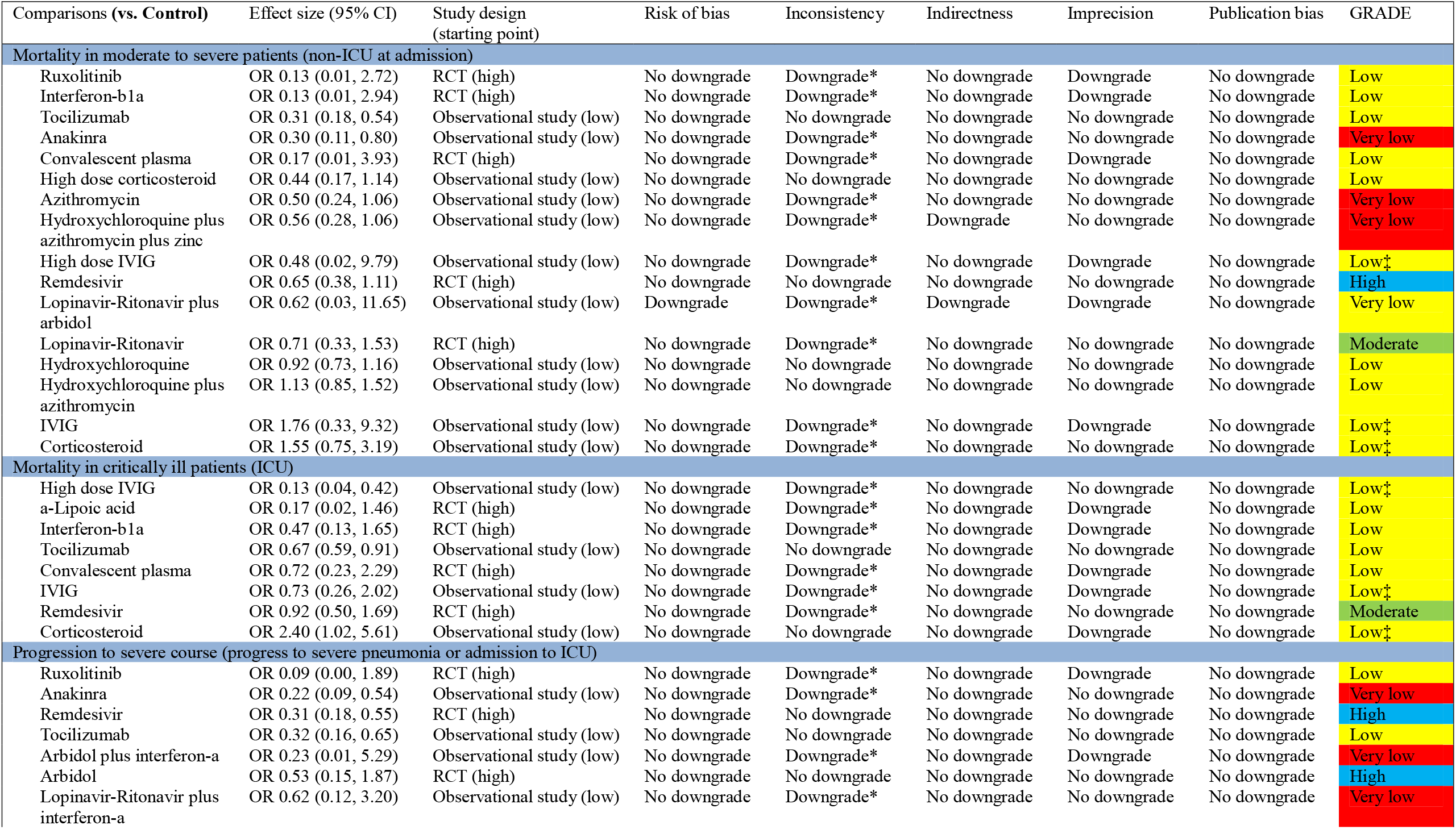
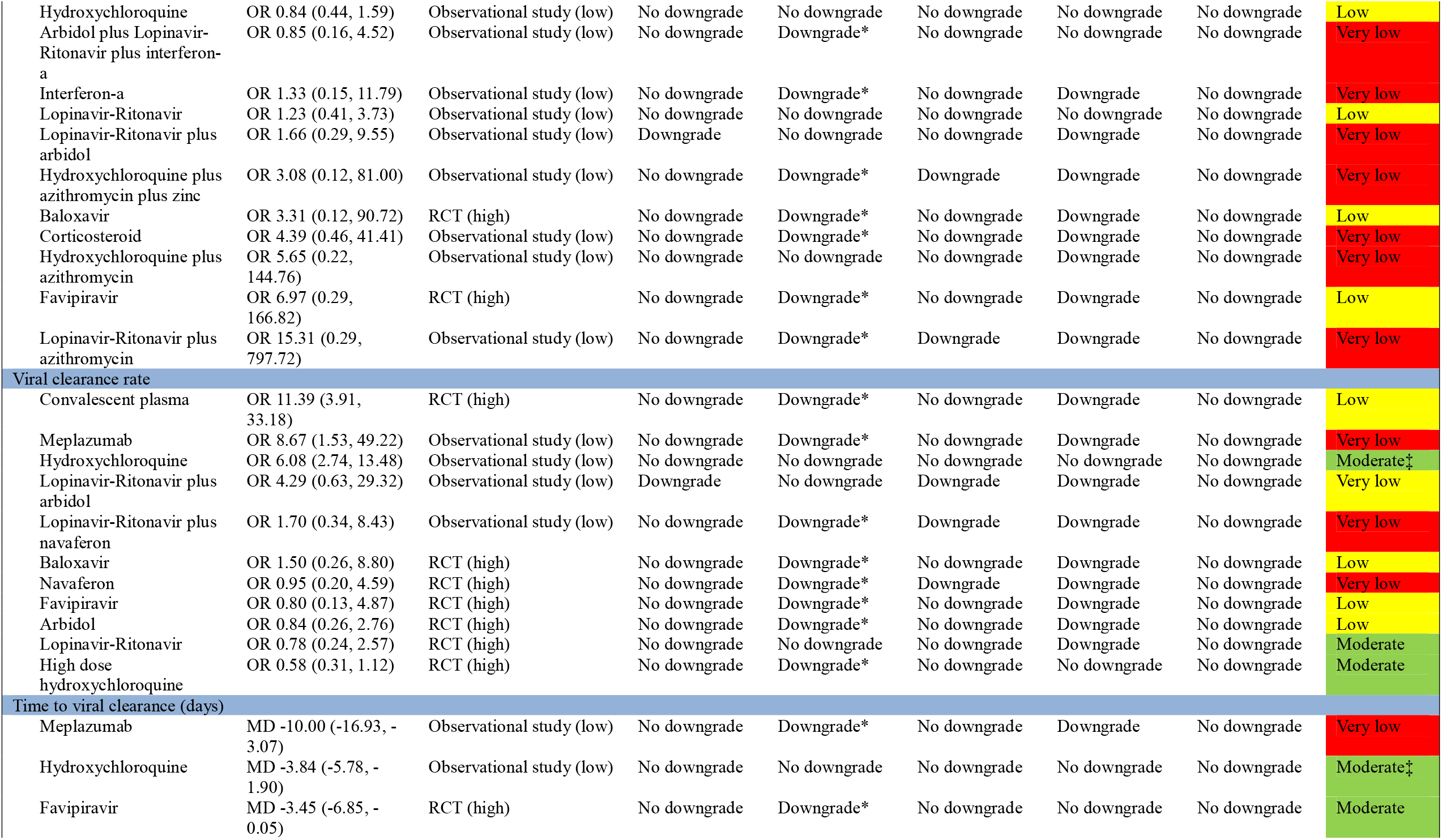
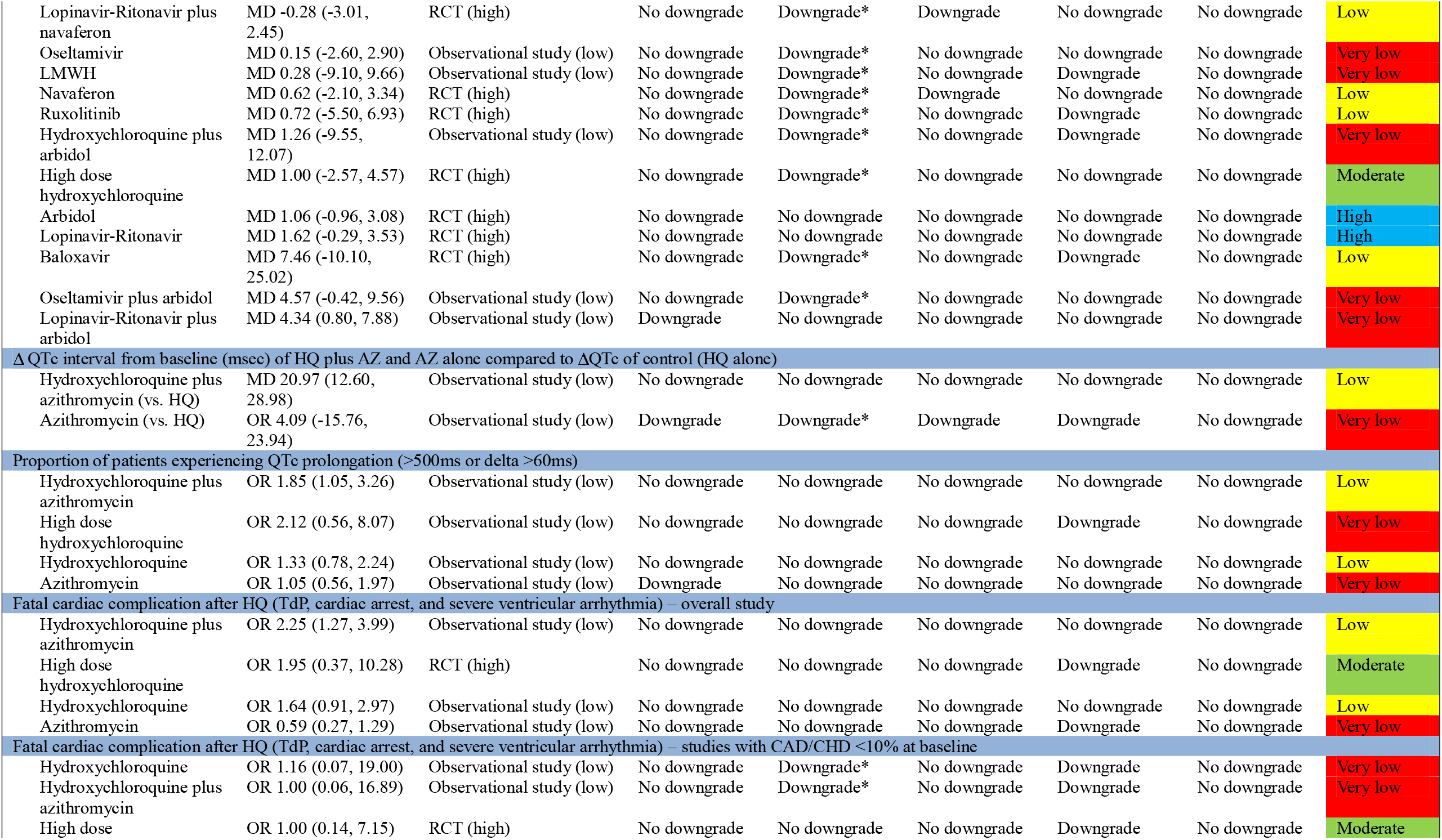
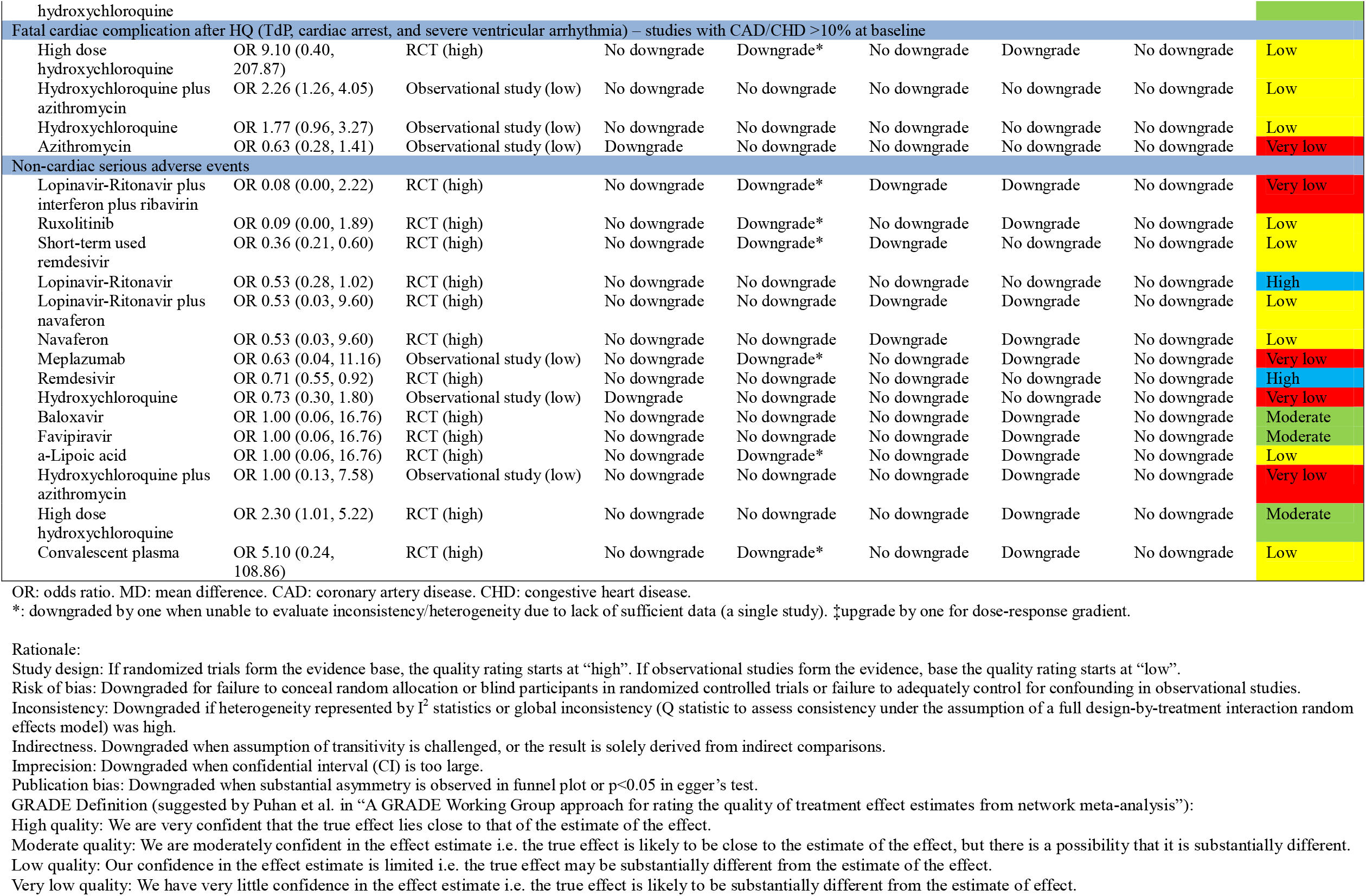
Certainty of evidence evaluated with Grading of Recommendations Assessment, Development, and Evaluation (GRADE) framework

| Comparisons (vs. Control) | Effect size (95% CI) | Study design (starting point) | Risk of bias | Inconsistency | Indirectness | Imprecision | Publication bias | GRADE |
| --- | --- | --- | --- | --- | --- | --- | --- | --- |
| **Mortality in moderate to severe patients (non-ICU at admission)** |  |  |  |  |  |  |  |  |
| Ruxolitinib | OR 0.13 (0.01, 2.72) | RCT (high) | No downgrade | Downgrade* | No downgrade | Downgrade | No downgrade | Low |
| Interferon-β1a | OR 0.13 (0.01, 2.94) | RCT (high) | No downgrade | Downgrade* | No downgrade | Downgrade | No downgrade | Low |
| Tocilizumab | OR 0.31 (0.18, 0.54) | Observational study (low) | No downgrade | No downgrade | No downgrade | No downgrade | No downgrade | Low |
| Anakinra | OR 0.30 (0.11, 0.80) | Observational study (low) | No downgrade | Downgrade* | No downgrade | No downgrade | No downgrade | Very low |
| Convalescent plasma | OR 0.17 (0.01, 3.93) | RCT (high) | No downgrade | Downgrade* | No downgrade | Downgrade | No downgrade | Low |
| High dose corticosteroid | OR 0.44 (0.17, 1.14) | Observational study (low) | No downgrade | No downgrade | No downgrade | No downgrade | No downgrade | Low |
| Azithromycin | OR 0.50 (0.24, 1.06) | Observational study (low) | No downgrade | Downgrade* | No downgrade | No downgrade | No downgrade | Very low |
| Hydroxychloroquine plus azithromycin plus zinc | OR 0.56 (0.28, 1.06) | Observational study (low) | No downgrade | Downgrade* | Downgrade | No downgrade | No downgrade | Very low |
| High dose IVIG | OR 0.48 (0.02, 9.79) | Observational study (low) | No downgrade | Downgrade* | No downgrade | Downgrade | No downgrade | Low† |
| Remdesivir | OR 0.65 (0.38, 1.11) | RCT (high) | No downgrade | No downgrade | No downgrade | No downgrade | No downgrade | High |
| Lopinavir-Ritonavir plus arbidol | OR 0.62 (0.03, 11.65) | Observational study (low) | Downgrade | Downgrade* | Downgrade | Downgrade | No downgrade | Very low |
| Lopinavir-Ritonavir | OR 0.71 (0.33, 1.53) | RCT (high) | No downgrade | Downgrade* | No downgrade | No downgrade | No downgrade | Moderate |
| Hydroxychloroquine | OR 0.92 (0.73, 1.16) | Observational study (low) | No downgrade | No downgrade | No downgrade | No downgrade | No downgrade | Low |
| Hydroxychloroquine plus azithromycin | OR 1.13 (0.85, 1.52) | Observational study (low) | No downgrade | No downgrade | No downgrade | No downgrade | No downgrade | Low |
| IVIG | OR 1.76 (0.33, 9.32) | Observational study (low) | No downgrade | Downgrade* | No downgrade | Downgrade | No downgrade | Low† |
| Corticosteroid | OR 1.55 (0.75, 3.19) | Observational study (low) | No downgrade | Downgrade* | No downgrade | No downgrade | No downgrade | Low† |
| **Mortality in critically ill patients (ICU)** |  |  |  |  |  |  |  |  |
| High dose IVIG | OR 0.13 (0.04, 0.42) | Observational study (low) | No downgrade | Downgrade* | No downgrade | No downgrade | No downgrade | Low† |
| a-Lipoic acid | OR 0.17 (0.02, 1.46) | RCT (high) | No downgrade | Downgrade* | No downgrade | Downgrade | No downgrade | Low |
| Interferon-β1a | OR 0.47 (0.13, 1.65) | RCT (high) | No downgrade | Downgrade* | No downgrade | Downgrade | No downgrade | Low |
| Tocilizumab | OR 0.67 (0.59, 0.91) | Observational study (low) | No downgrade | No downgrade | No downgrade | No downgrade | No downgrade | Low |
| Convalescent plasma | OR 0.72 (0.23, 2.29) | RCT (high) | No downgrade | Downgrade* | No downgrade | Downgrade | No downgrade | Low |
| IVIG | OR 0.73 (0.26, 2.02) | Observational study (low) | No downgrade | Downgrade* | No downgrade | Downgrade | No downgrade | Low† |
| Remdesivir | OR 0.92 (0.50, 1.69) | RCT (high) | No downgrade | Downgrade* | No downgrade | No downgrade | No downgrade | Moderate |
| Corticosteroid | OR 2.40 (1.02, 5.61) | Observational study (low) | No downgrade | No downgrade | No downgrade | Downgrade | No downgrade | Low† |
| **Progression to severe course (progress to severe pneumonia or admission to ICU)** |  |  |  |  |  |  |  |  |
| Ruxolitinib | OR 0.09 (0.00, 1.89) | RCT (high) | No downgrade | Downgrade* | No downgrade | Downgrade | No downgrade | Low |
| Anakinra | OR 0.22 (0.09, 0.54) | Observational study (low) | No downgrade | Downgrade* | No downgrade | No downgrade | No downgrade | Very low |
| Remdesivir | OR 0.31 (0.18, 0.55) | RCT (high) | No downgrade | No downgrade | No downgrade | No downgrade | No downgrade | High |
| Tocilizumab | OR 0.32 (0.16, 0.65) | Observational study (low) | No downgrade | No downgrade | No downgrade | No downgrade | No downgrade | Low |
| Arbidol plus interferon-a | OR 0.23 (0.01, 5.29) | Observational study (low) | No downgrade | Downgrade* | No downgrade | Downgrade | No downgrade | Very low |
| Arbidol | OR 0.53 (0.15, 1.87) | RCT (high) | No downgrade | No downgrade | No downgrade | No downgrade | No downgrade | High |
| Lopinavir-Ritonavir plus interferon-a | OR 0.62 (0.12, 3.20) | Observational study (low) | No downgrade | Downgrade* | No downgrade | No downgrade | No downgrade | Very low |
| Hydroxychloroquine | OR 0.84 (0.44, 1.59) | Observational study (low) | No downgrade | No downgrade | No downgrade | No downgrade | No downgrade | Low |
| Arbidol plus Lopinavir-Ritonavir plus interferon-a | OR 0.85 (0.16, 4.52) | Observational study (low) | No downgrade | Downgrade* | No downgrade | No downgrade | No downgrade | Very low |
| Interferon-a | OR 1.33 (0.15, 11.79) | Observational study (low) | No downgrade | Downgrade* | No downgrade | Downgrade | No downgrade | Very low |
| Lopinavir-Ritonavir | OR 1.23 (0.41, 3.73) | Observational study (low) | No downgrade | No downgrade | No downgrade | No downgrade | No downgrade | Low |
| Lopinavir-Ritonavir plus arbidol | OR 1.66 (0.29, 9.55) | Observational study (low) | Downgrade | No downgrade | No downgrade | Downgrade | No downgrade | Very low |
| Hydroxychloroquine plus azithromycin plus zinc | OR 3.08 (0.12, 81.00) | Observational study (low) | No downgrade | Downgrade* | Downgrade | Downgrade | No downgrade | Very low |
| Baloxavir | OR 3.31 (0.12, 90.72) | RCT (high) | No downgrade | Downgrade* | No downgrade | Downgrade | No downgrade | Low |
| Corticosteroid | OR 4.39 (0.46, 41.41) | Observational study (low) | No downgrade | Downgrade* | No downgrade | Downgrade | No downgrade | Very low |
| Hydroxychloroquine plus azithromycin | OR 5.65 (0.22, 144.76) | Observational study (low) | No downgrade | No downgrade | No downgrade | Downgrade | No downgrade | Very low |
| Favipiravir | OR 6.97 (0.29, 166.82) | RCT (high) | No downgrade | Downgrade* | No downgrade | Downgrade | No downgrade | Low |
| Lopinavir-Ritonavir plus azithromycin | OR 15.31 (0.29, 797.72) | Observational study (low) | No downgrade | Downgrade* | Downgrade | Downgrade | No downgrade | Very low |
| Viral clearance rate |  |  |  |  |  |  |  |  |
| Convalescent plasma | OR 11.39 (3.91, 33.18) | RCT (high) | No downgrade | Downgrade* | No downgrade | Downgrade | No downgrade | Low |
| Meplazumab | OR 8.67 (1.53, 49.22) | Observational study (low) | No downgrade | Downgrade* | No downgrade | Downgrade | No downgrade | Very low |
| Hydroxychloroquine | OR 6.08 (2.74, 13.48) | Observational study (low) | No downgrade | No downgrade | No downgrade | No downgrade | No downgrade | Moderate‡ |
| Lopinavir-Ritonavir plus arbidol | OR 4.29 (0.63, 29.32) | Observational study (low) | Downgrade | No downgrade | Downgrade | Downgrade | No downgrade | Very low |
| Lopinavir-Ritonavir plus navaferon | OR 1.70 (0.34, 8.43) | Observational study (low) | No downgrade | Downgrade* | Downgrade | Downgrade | No downgrade | Very low |
| Baloxavir | OR 1.50 (0.26, 8.80) | RCT (high) | No downgrade | Downgrade* | No downgrade | Downgrade | No downgrade | Low |
| Navaferon | OR 0.95 (0.20, 4.59) | RCT (high) | No downgrade | Downgrade* | Downgrade | Downgrade | No downgrade | Very low |
| Favipiravir | OR 0.80 (0.13, 4.87) | RCT (high) | No downgrade | Downgrade* | No downgrade | Downgrade | No downgrade | Low |
| Arbidol | OR 0.84 (0.26, 2.76) | RCT (high) | No downgrade | Downgrade* | No downgrade | Downgrade | No downgrade | Low |
| Lopinavir-Ritonavir | OR 0.78 (0.24, 2.57) | RCT (high) | No downgrade | No downgrade | No downgrade | Downgrade | No downgrade | Moderate |
| High dose hydroxychloroquine | OR 0.58 (0.31, 1.12) | RCT (high) | No downgrade | Downgrade* | No downgrade | No downgrade | No downgrade | Moderate |
| Time to viral clearance (days) |  |  |  |  |  |  |  |  |
| Meplazumab | MD -10.00 (-16.93, -3.07) | Observational study (low) | No downgrade | Downgrade* | No downgrade | Downgrade | No downgrade | Very low |
| Hydroxychloroquine | MD -3.84 (-5.78, -1.90) | Observational study (low) | No downgrade | No downgrade | No downgrade | No downgrade | No downgrade | Moderate‡ |
| Favipiravir | MD -3.45 (-6.85, -0.05) | RCT (high) | No downgrade | Downgrade* | No downgrade | No downgrade | No downgrade | Moderate |
| Lopinavir-Ritonavir plus navaferon | MD -0.28 (-3.01, 2.45) | RCT (high) | No downgrade | Downgrade* | Downgrade | No downgrade | No downgrade | Low |
| Oseltamivir | MD 0.15 (-2.60, 2.90) | Observational study (low) | No downgrade | Downgrade* | No downgrade | No downgrade | No downgrade | Very low |
| LMWH | MD 0.28 (-9.10, 9.66) | Observational study (low) | No downgrade | Downgrade* | No downgrade | Downgrade | No downgrade | Very low |
| Navaferon | MD 0.62 (-2.10, 3.34) | RCT (high) | No downgrade | Downgrade* | Downgrade | No downgrade | No downgrade | Low |
| Ruxolitinib | MD 0.72 (-5.50, 6.93) | RCT (high) | No downgrade | Downgrade* | No downgrade | Downgrade | No downgrade | Low |
| Hydroxychloroquine plus arbidol | MD 1.26 (-9.55, 12.07) | Observational study (low) | No downgrade | Downgrade* | No downgrade | Downgrade | No downgrade | Very low |
| High dose hydroxychloroquine | MD 1.00 (-2.57, 4.57) | RCT (high) | No downgrade | Downgrade* | No downgrade | No downgrade | No downgrade | Moderate |
| Arbidol | MD 1.06 (-0.96, 3.08) | RCT (high) | No downgrade | No downgrade | No downgrade | No downgrade | No downgrade | High |
| Lopinavir-Ritonavir | MD 1.62 (-0.29, 3.53) | RCT (high) | No downgrade | No downgrade | No downgrade | No downgrade | No downgrade | High |
| Baloxavir | MD 7.46 (-10.10, 25.02) | RCT (high) | No downgrade | Downgrade* | No downgrade | Downgrade | No downgrade | Low |
| Oseltamivir plus arbidol | MD 4.57 (-0.42, 9.56) | Observational study (low) | No downgrade | Downgrade* | No downgrade | No downgrade | No downgrade | Very low |
| Lopinavir-Ritonavir plus arbidol | MD 4.34 (0.80, 7.88) | Observational study (low) | Downgrade | No downgrade | No downgrade | No downgrade | No downgrade | Very low |
| **Δ QTc interval from baseline (msec) of HQ plus AZ and AZ alone compared to ΔQTc of control (HQ alone)** |  |  |  |  |  |  |  |  |
| Hydroxychloroquine plus azithromycin (vs. HQ) | MD 20.97 (12.60, 28.98) | Observational study (low) | No downgrade | No downgrade | No downgrade | No downgrade | No downgrade | Low |
| Azithromycin (vs. HQ) | OR 4.09 (-15.76, 23.94) | Observational study (low) | Downgrade | Downgrade* | Downgrade | Downgrade | No downgrade | Very low |
| **Proportion of patients experiencing QTc prolongation (>500ms or delta >60ms)** |  |  |  |  |  |  |  |  |
| Hydroxychloroquine plus azithromycin | OR 1.85 (1.05, 3.26) | Observational study (low) | No downgrade | No downgrade | No downgrade | No downgrade | No downgrade | Low |
| High dose hydroxychloroquine | OR 2.12 (0.56, 8.07) | Observational study (low) | No downgrade | No downgrade | No downgrade | Downgrade | No downgrade | Very low |
| Hydroxychloroquine | OR 1.33 (0.78, 2.24) | Observational study (low) | No downgrade | No downgrade | No downgrade | No downgrade | No downgrade | Low |
| Azithromycin | OR 1.05 (0.56, 1.97) | Observational study (low) | Downgrade | No downgrade | No downgrade | No downgrade | No downgrade | Very low |
| **Fatal cardiac complication after HQ (TdP, cardiac arrest, and severe ventricular arrhythmia) – overall study** |  |  |  |  |  |  |  |  |
| Hydroxychloroquine plus azithromycin | OR 2.25 (1.27, 3.99) | Observational study (low) | No downgrade | No downgrade | No downgrade | No downgrade | No downgrade | Low |
| High dose hydroxychloroquine | OR 1.95 (0.37, 10.28) | RCT (high) | No downgrade | No downgrade | No downgrade | Downgrade | No downgrade | Moderate |
| Hydroxychloroquine | OR 1.64 (0.91, 2.97) | Observational study (low) | No downgrade | No downgrade | No downgrade | No downgrade | No downgrade | Low |
| Azithromycin | OR 0.59 (0.27, 1.29) | Observational study (low) | No downgrade | No downgrade | No downgrade | Downgrade | No downgrade | Very low |
| **Fatal cardiac complication after HQ (TdP, cardiac arrest, and severe ventricular arrhythmia) – studies with CAD/CHD <10% at baseline** |  |  |  |  |  |  |  |  |
| Hydroxychloroquine | OR 1.16 (0.07, 19.00) | Observational study (low) | No downgrade | Downgrade* | No downgrade | Downgrade | No downgrade | Very low |
| Hydroxychloroquine plus azithromycin | OR 1.00 (0.06, 16.89) | Observational study (low) | No downgrade | Downgrade* | No downgrade | Downgrade | No downgrade | Very low |
| High dose | OR 1.00 (0.14, 7.15) | RCT (high) | No downgrade | No downgrade | No downgrade | Downgrade | No downgrade | Moderate |
| hydroxychloroquine |  |  |  |  |  |  |  |  |
| Fatal cardiac complication after HQ (TdP, cardiac arrest, and severe ventricular arrhythmia) – studies with CAD/CHD >10% at baseline |  |  |  |  |  |  |  |  |
| High dose hydroxychloroquine | OR 9.10 (0.40, 207.87) | RCT (high) | No downgrade | Downgrade* | No downgrade | Downgrade | No downgrade | Low |
| Hydroxychloroquine plus azithromycin | OR 2.26 (1.26, 4.05) | Observational study (low) | No downgrade | No downgrade | No downgrade | No downgrade | No downgrade | Low |
| Hydroxychloroquine | OR 1.77 (0.96, 3.27) | Observational study (low) | No downgrade | No downgrade | No downgrade | No downgrade | No downgrade | Low |
| Azithromycin | OR 0.63 (0.28, 1.41) | Observational study (low) | Downgrade | No downgrade | No downgrade | No downgrade | No downgrade | Very low |
| Non-cardiac serious adverse events |  |  |  |  |  |  |  |  |
| Lopinavir-Ritonavir plus interferon plus ribavirin | OR 0.08 (0.00, 2.22) | RCT (high) | No downgrade | Downgrade* | Downgrade | Downgrade | No downgrade | Very low |
| Ruxolitinib | OR 0.09 (0.00, 1.89) | RCT (high) | No downgrade | Downgrade* | No downgrade | Downgrade | No downgrade | Low |
| Short-term used remdesivir | OR 0.36 (0.21, 0.60) | RCT (high) | No downgrade | Downgrade* | Downgrade | No downgrade | No downgrade | Low |
| Lopinavir-Ritonavir | OR 0.53 (0.28, 1.02) | RCT (high) | No downgrade | No downgrade | No downgrade | No downgrade | No downgrade | High |
| Lopinavir-Ritonavir plus navaferon | OR 0.53 (0.03, 9.60) | RCT (high) | No downgrade | No downgrade | Downgrade | Downgrade | No downgrade | Low |
| Navaferon | OR 0.53 (0.03, 9.60) | RCT (high) | No downgrade | No downgrade | Downgrade | Downgrade | No downgrade | Low |
| Meplazumab | OR 0.63 (0.04, 11.16) | Observational study (low) | No downgrade | Downgrade* | No downgrade | Downgrade | No downgrade | Very low |
| Remdesivir | OR 0.71 (0.55, 0.92) | RCT (high) | No downgrade | No downgrade | No downgrade | No downgrade | No downgrade | High |
| Hydroxychloroquine | OR 0.73 (0.30, 1.80) | Observational study (low) | Downgrade | No downgrade | No downgrade | No downgrade | No downgrade | Very low |
| Baloxavir | OR 1.00 (0.06, 16.76) | RCT (high) | No downgrade | No downgrade | No downgrade | Downgrade | No downgrade | Moderate |
| Favipiravir | OR 1.00 (0.06, 16.76) | RCT (high) | No downgrade | No downgrade | No downgrade | Downgrade | No downgrade | Moderate |
| a-Lipoic acid | OR 1.00 (0.06, 16.76) | RCT (high) | No downgrade | Downgrade* | No downgrade | Downgrade | No downgrade | Low |
| Hydroxychloroquine plus azithromycin | OR 1.00 (0.13, 7.58) | Observational study (low) | No downgrade | No downgrade | No downgrade | Downgrade | No downgrade | Very low |
| High dose hydroxychloroquine | OR 2.30 (1.01, 5.22) | RCT (high) | No downgrade | No downgrade | No downgrade | Downgrade | No downgrade | Moderate |
| Convalescent plasma | OR 5.10 (0.24, 108.86) | RCT (high) | No downgrade | Downgrade* | No downgrade | Downgrade | No downgrade | Low |

OR: odds ratio. MD: mean difference. CAD: coronary artery disease. CHD: congestive heart disease.
\*: downgraded by one when unable to evaluate inconsistency/heterogeneity due to lack of sufficient data (a single study). ‡upgrade by one for dose-response gradient.

**Figure 2:**
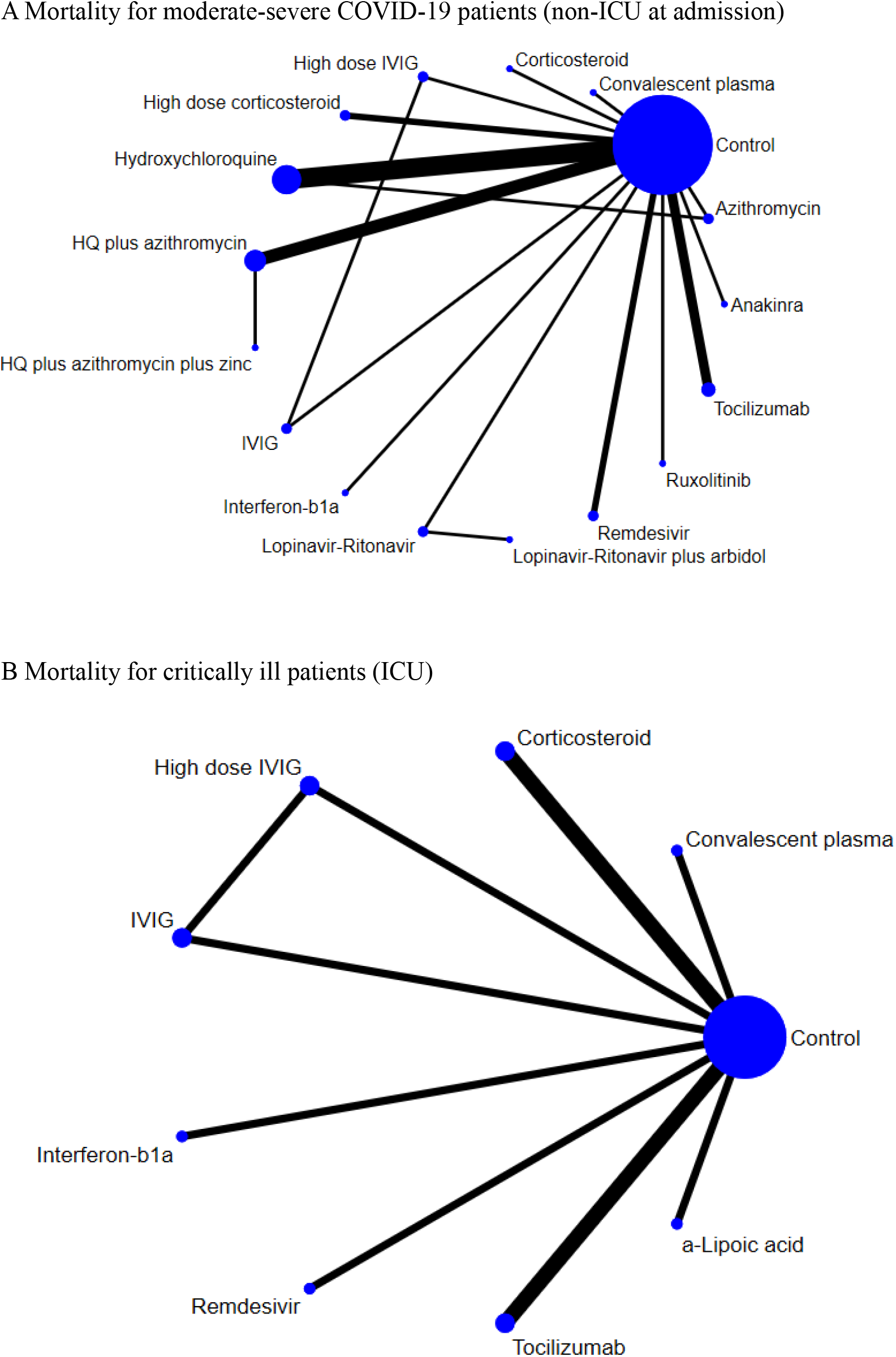
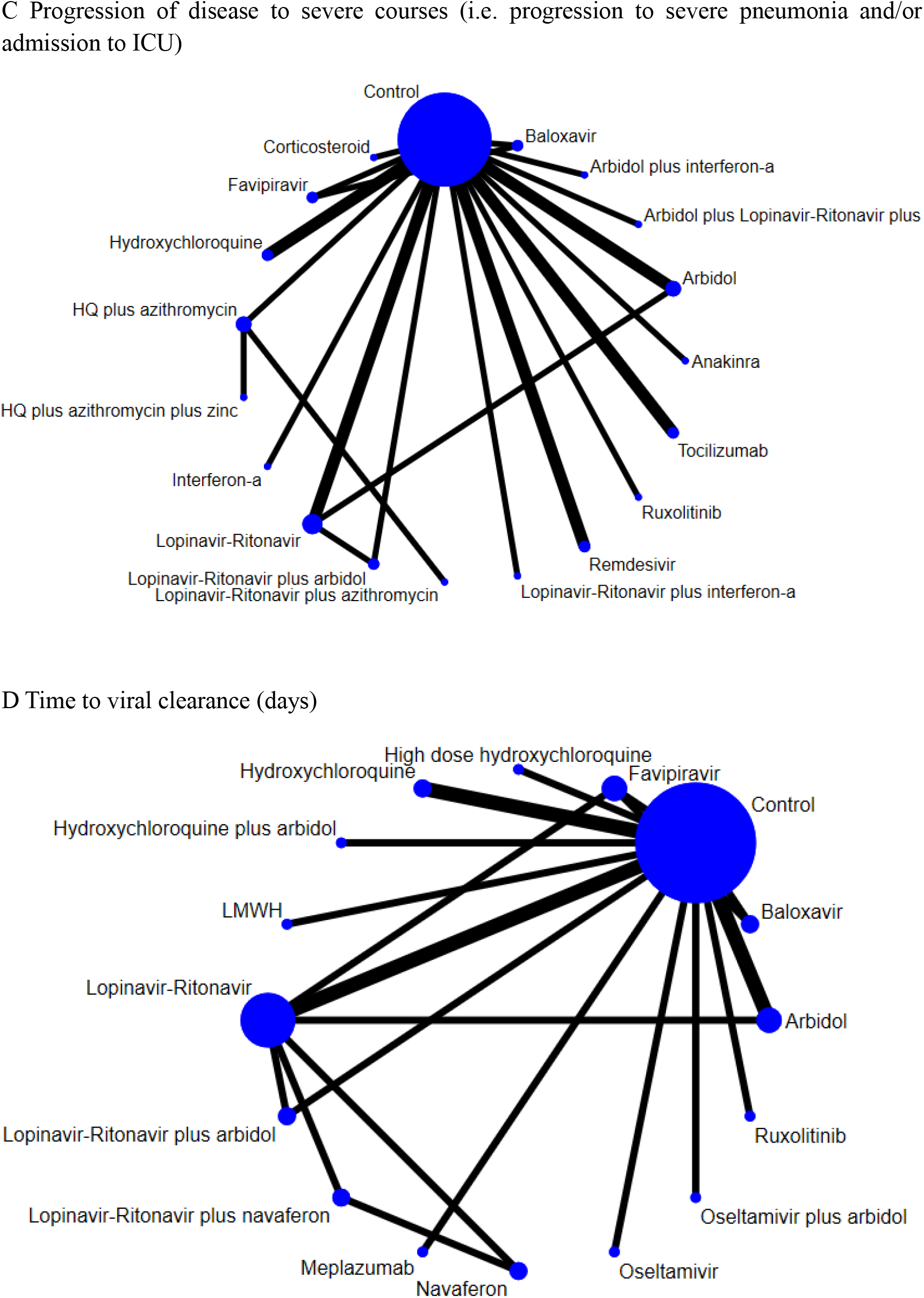
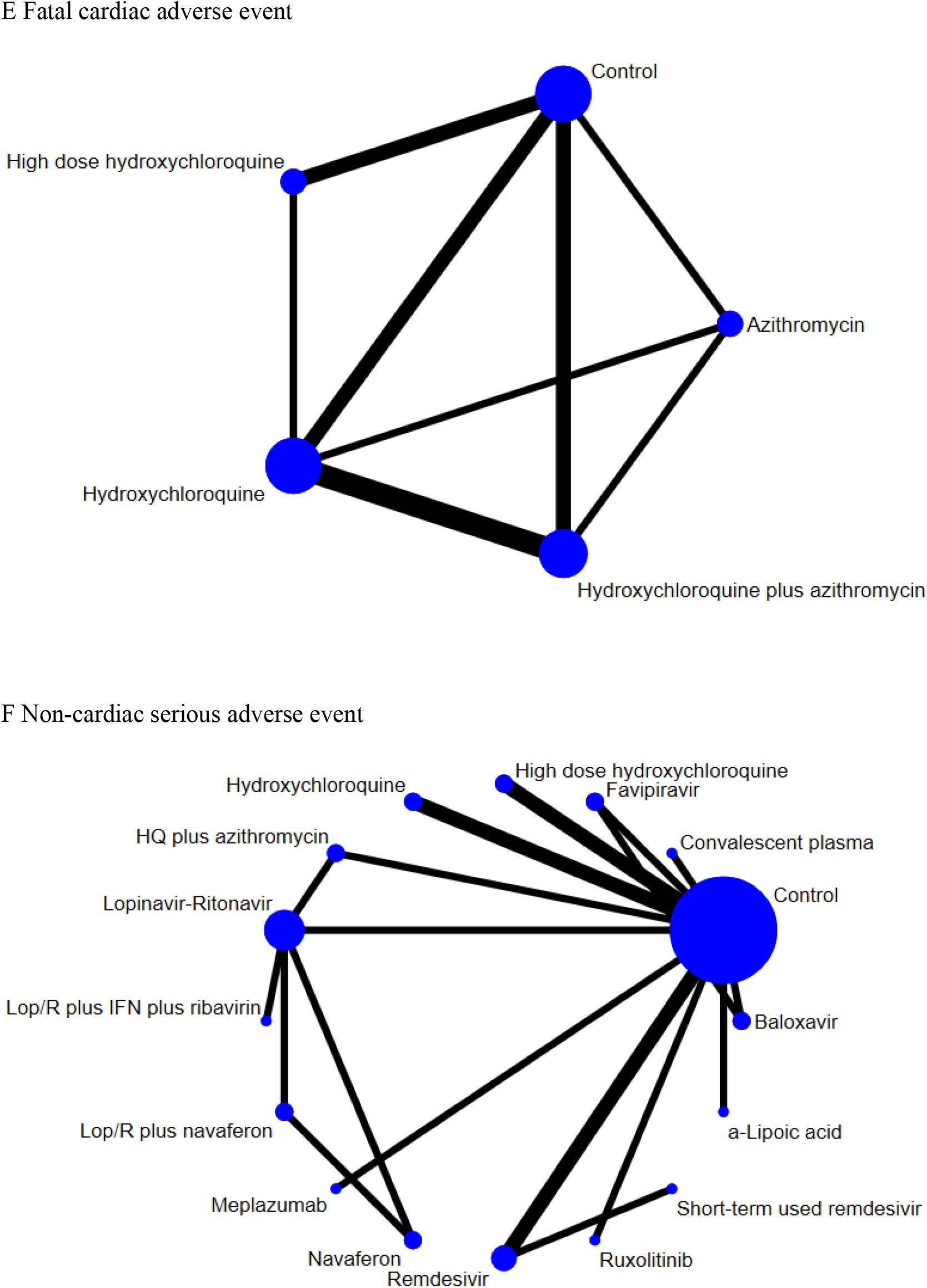
Network of eligible comparisons for primary outcomes (A) Mortality for moderate-severe COVID-19 patients (non-ICU at admission). (B) mortality for critically ill patients (ICU). (C) Progression of disease to severe courses (i.e. progression to severe pneumonia and/or admission to ICU). (D) Time to viral clearance (days). (E) Fatal cardiac adverse events (torsades de pointes, cardiac arrest, and severe ventricular arrhythmia). (F) Non-cardiac serious adverse events. Lines indicate direct comparison of agents, and the thickness of line corresponds to the number of trials in the comparison. Size of node corresponds to the number of studies that involve the intervention. HQ = Hydroxychloroquine. Lop/R = Lopinavir-Ritonavir. ICU = intensive care unit.

### Mortality in ICU and non-ICU settings

Tocilizumab (Odds ratio (OR) 0.31, 95% confidence interval (CI) 0.18 to 0.54, low certainty) and anakinra (OR 0.30, 95% CI 0.11 to 0.80, very low certainty) significantly reduced the mortality in moderate-to-severe patients hospitalized in a non-ICU setting compared to the control group (Figure 3A). This effect could not be confirmed in a parallel sensitivity analysis with only RCTs (Figure 3B) as no RCTs were conducted for either agent. In critically ill patients hospitalized in the ICU, high dose IVIG (OR 0.13, 95% CI 0.04 to 0.42, low certainty) and tocilizumab (OR 0.67, 95% CI 0.50 to 0.91, low certainty) were shown to lower morality while corticosteroid therapy was shown to increase mortality (OR 2.40, 95% CI 1.02 to 5.61, moderate certainty) (Figure 3C).

**Figure 3:**
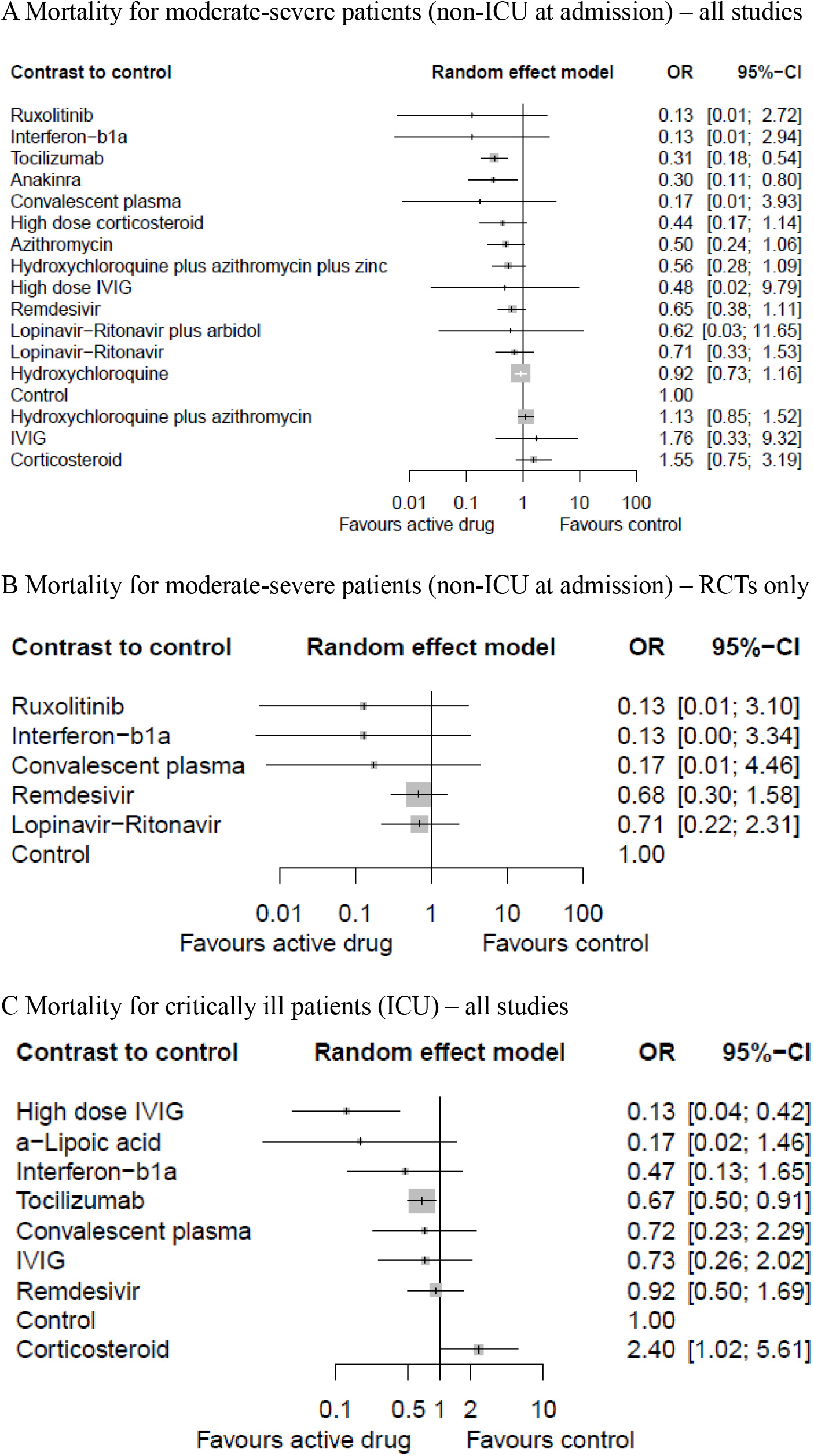
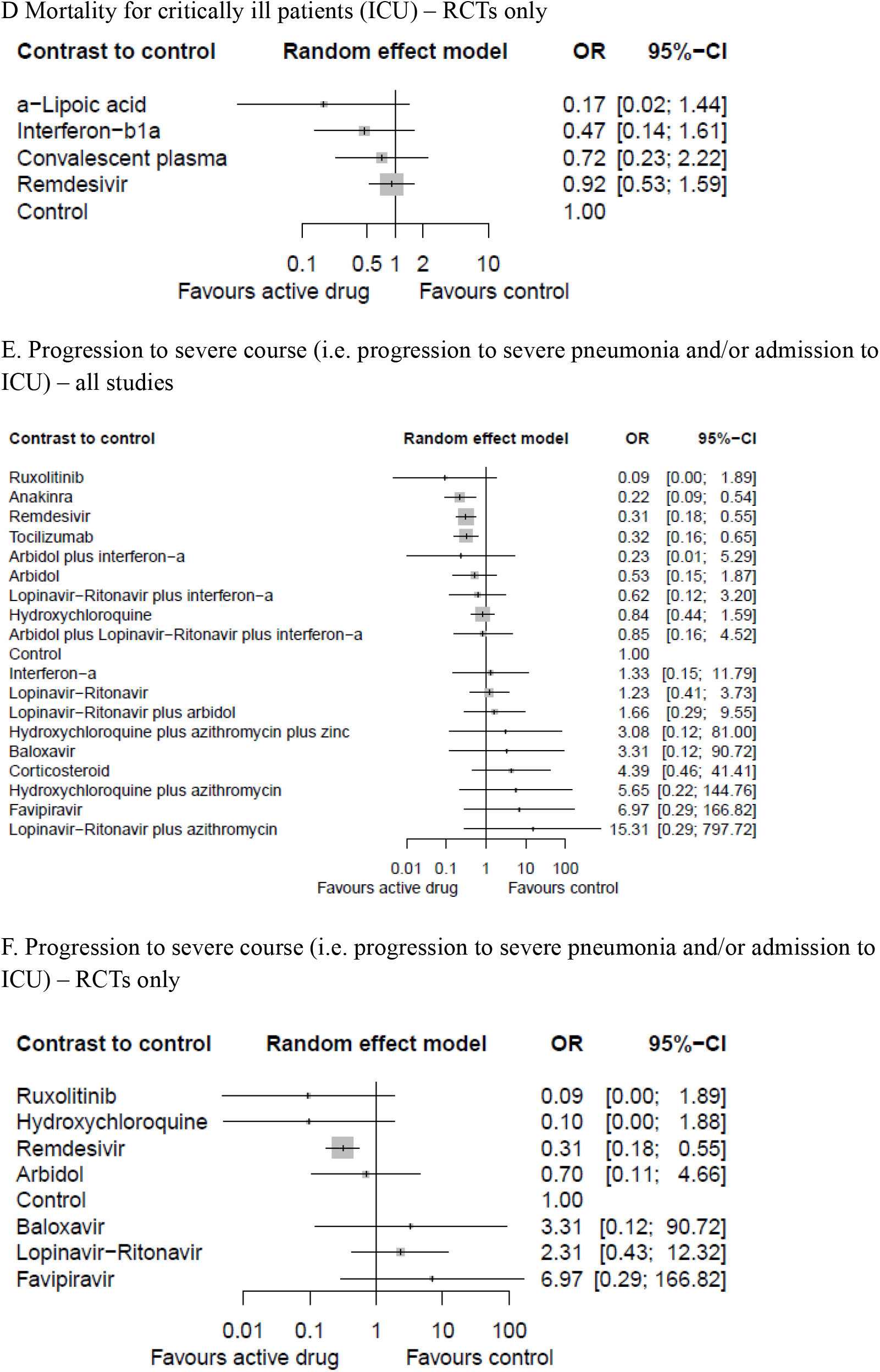
Network meta-analysis of pharmacological interventions compared with control (standard care) for efficacy outcomes. Mortality for moderate-severe patients (non-ICU at admission) from (A) all studies and (B) RCTs only. Mortality for critically ill patients (ICU) from (C) all studies and (D) RCTs only. Progression to severe course (i.e. progression to severe pneumonia and/or admission to ICU) from (E) all studies and (F) RCTs only. Effect estimates are presented in odds ratios (OR) with 95% CI. Pharmacological agents are ranked by surface under the cumulative ranking curve (SUCRA) value. RCT = randomized controlled trial. ICU = intensive care unit.

### Progression to severe pneumonia or admission to ICU

Tocilizumab (OR 0.32, 95% CI 0.16 to 065, low certainty), anakinra (OR 0.22, 95% CI 0.09 to 0.54, very low certainty), and remdesivir (OR 0.31, 95% CI 0.18 to 0.55, high certainty) showed effectiveness in preventing progression to severe courses (Figure 3E). Only remdesivir was shown to be effective in the analysis using only RCTs (OR 0.31, 95% CI 0.18 to 0.55) (Figure 3F).

### Viral clearance rate (negative conversion rate)

The use of convalescent plasma (OR 11.39, 95% CI 3.91 to 33.18, low certainty), hydroxychloroquine (OR 6.08, 95% CI 2.74 to 13.48, moderate certainty), and meplazumab (OR 8.67, 95% CI 1.53 to 49.22, very low certainty) showed significantly higher viral clearance rate compared to standard supportive therapy (Figure 4A). However, this effect of hydroxychloroquine and meplazumab was not replicated in the analysis of only RCTs (Figure 4B). The result was similar when using the continuous variable of time to viral clearance (days) as the outcome measure (Figure 4C, D).

**Figure 4:**
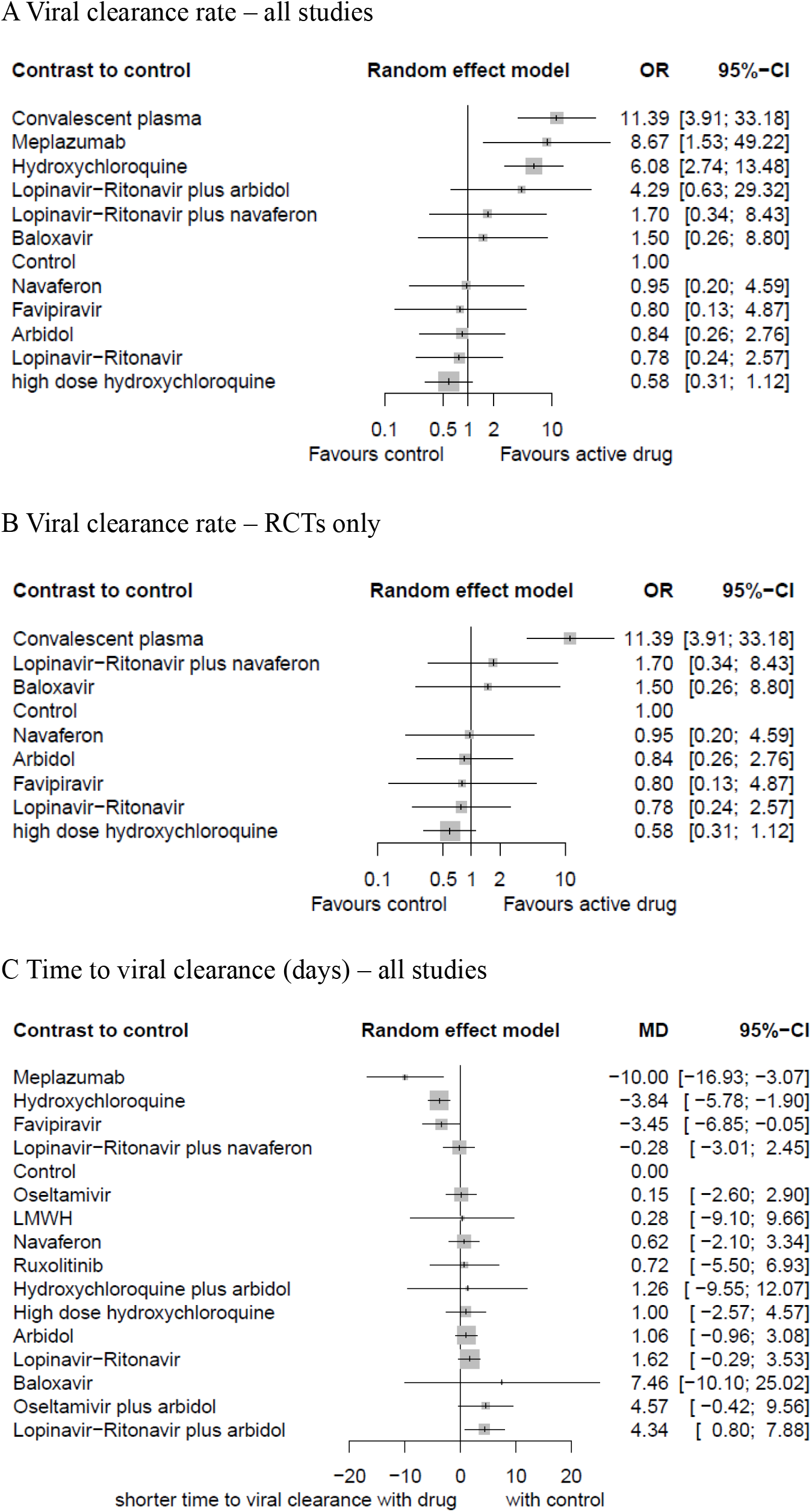
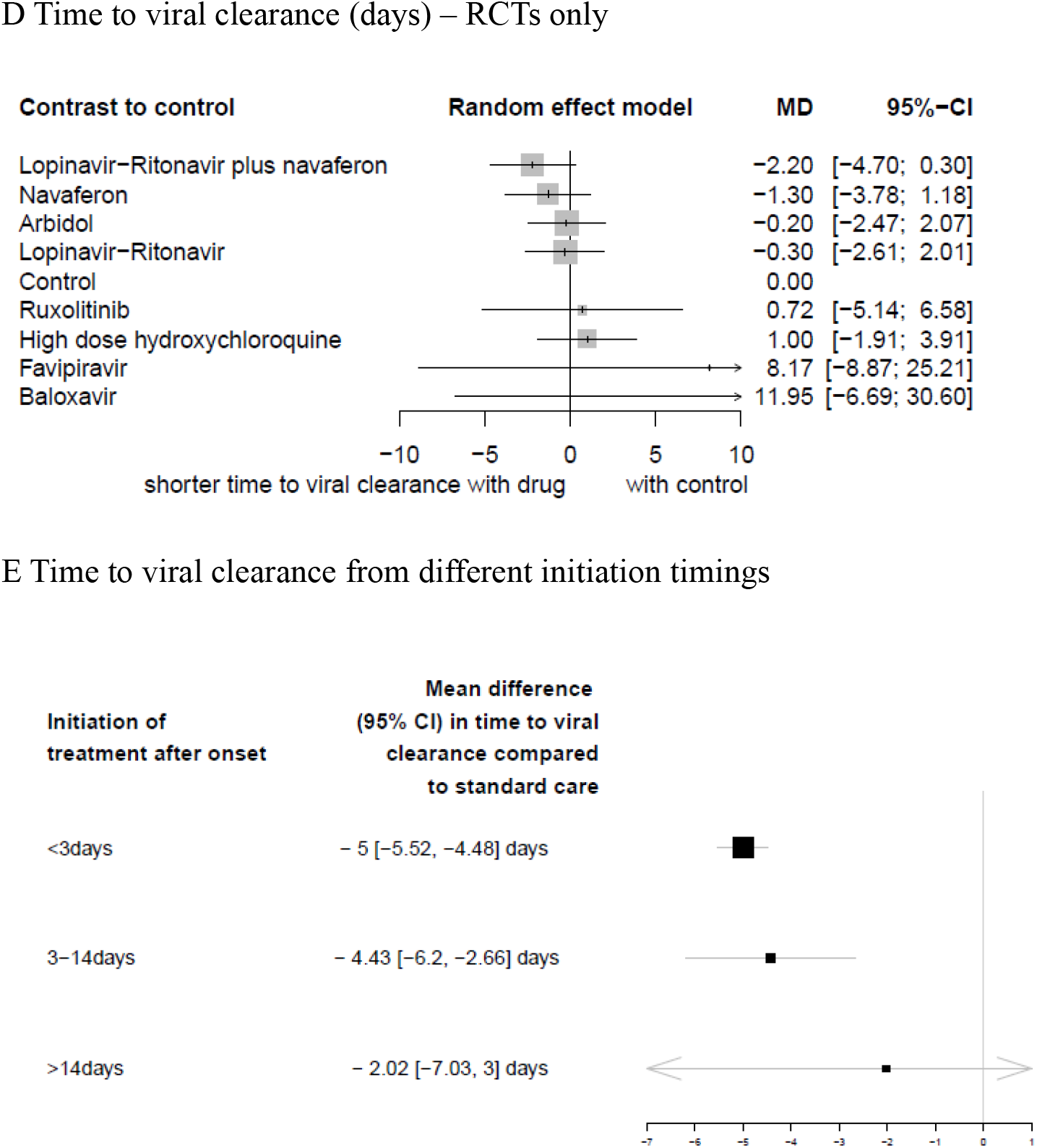
Network meta-analysis of pharmacological interventions compared with control (standard care) for viral clearance. Viral clearance rate (proportion of patients converted to PCR-negative status) from (A) all studies and (B) RCTs only. Time to viral clearance (days) from (C) all studies and (D) RCTs only. (E) Time to viral clearance from different hydroxychloroquine treatment initiation timings after symptom onset. Effect estimates are presented in odds ratios (OR) for viral clearance rate and mean differences (MD) for time to viral clearance, with 95% CI. Pharmacological agents are ranked by surface under the cumulative ranking curve (SUCRA) value. RCT = randomized controlled trial.

### Time to treatment initiation from symptom onset

The effect of the timing of hydroxychloroquine treatment initiation after the symptom onset (Figure 4E) was assessed. Treatment initiated after 14 days (MD −2.02, 95% CI −7.03 to 3.00) from symptom onset did not reduce the time to viral clearance compared to standard care.

### QTc prolongation

Compared to hydroxychloroquine monotherapy, the prolongation of QTc interval after treatment initiation was statistically significantly longer in the hydroxychloroquine plus azithromycin group (MD 20.79ms, 95% CI 12.60 to 28.98, low certainty) (Figure 5A). The proportion of patients experiencing QTc prolongation (defined by QTc interval >500ms or ΔQTc >60ms) was also significantly higher in the hydroxychloroquine plus azithromycin group compared to the control group (OR 1.85, 95% CI 1.05 to 3.26, very low certainty) but not in the hydroxychloroquine monotherapy group, azithromycin monotherapy group, or high-dose hydroxychloroquine group (Figure 5B).

**Figure 5:**
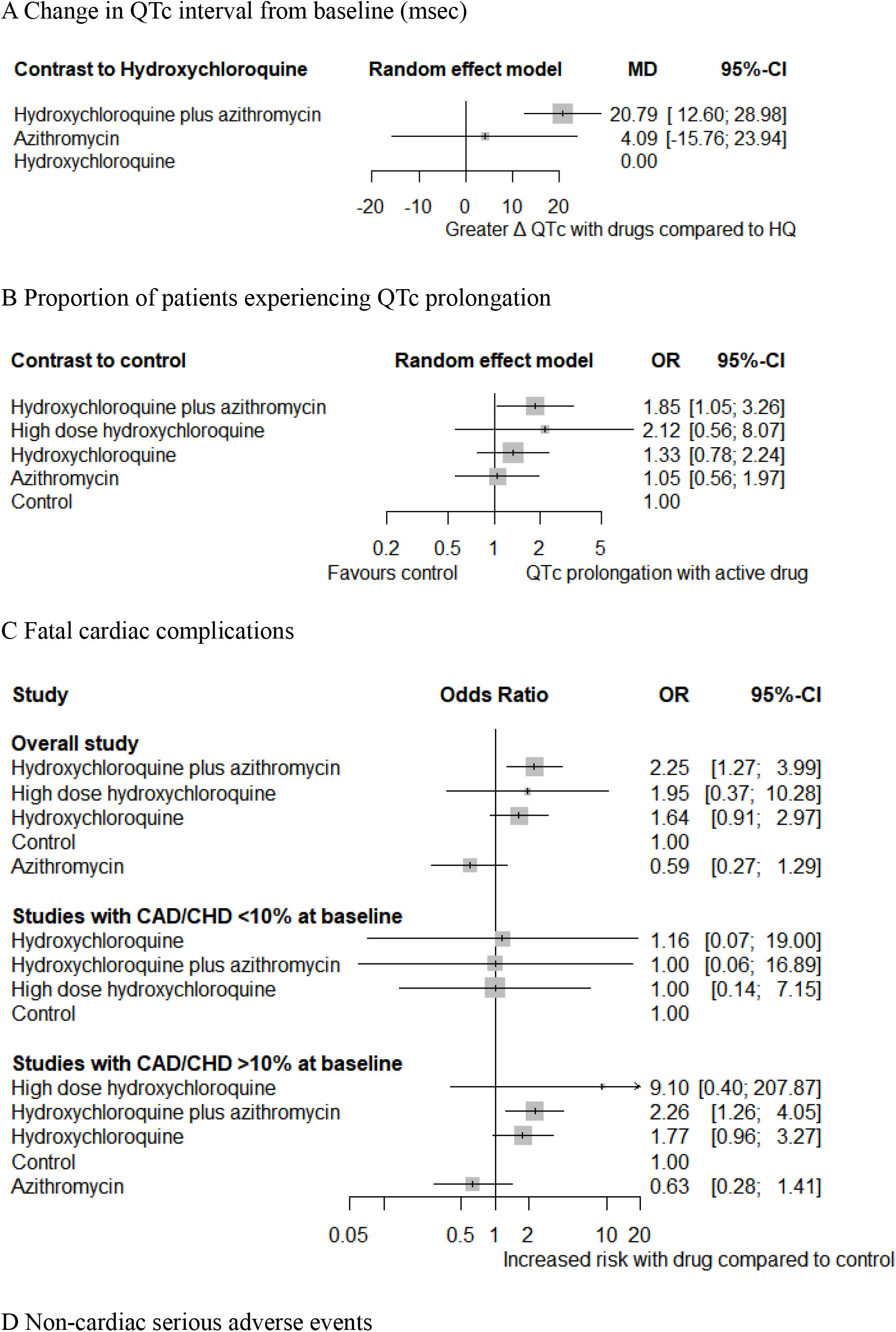
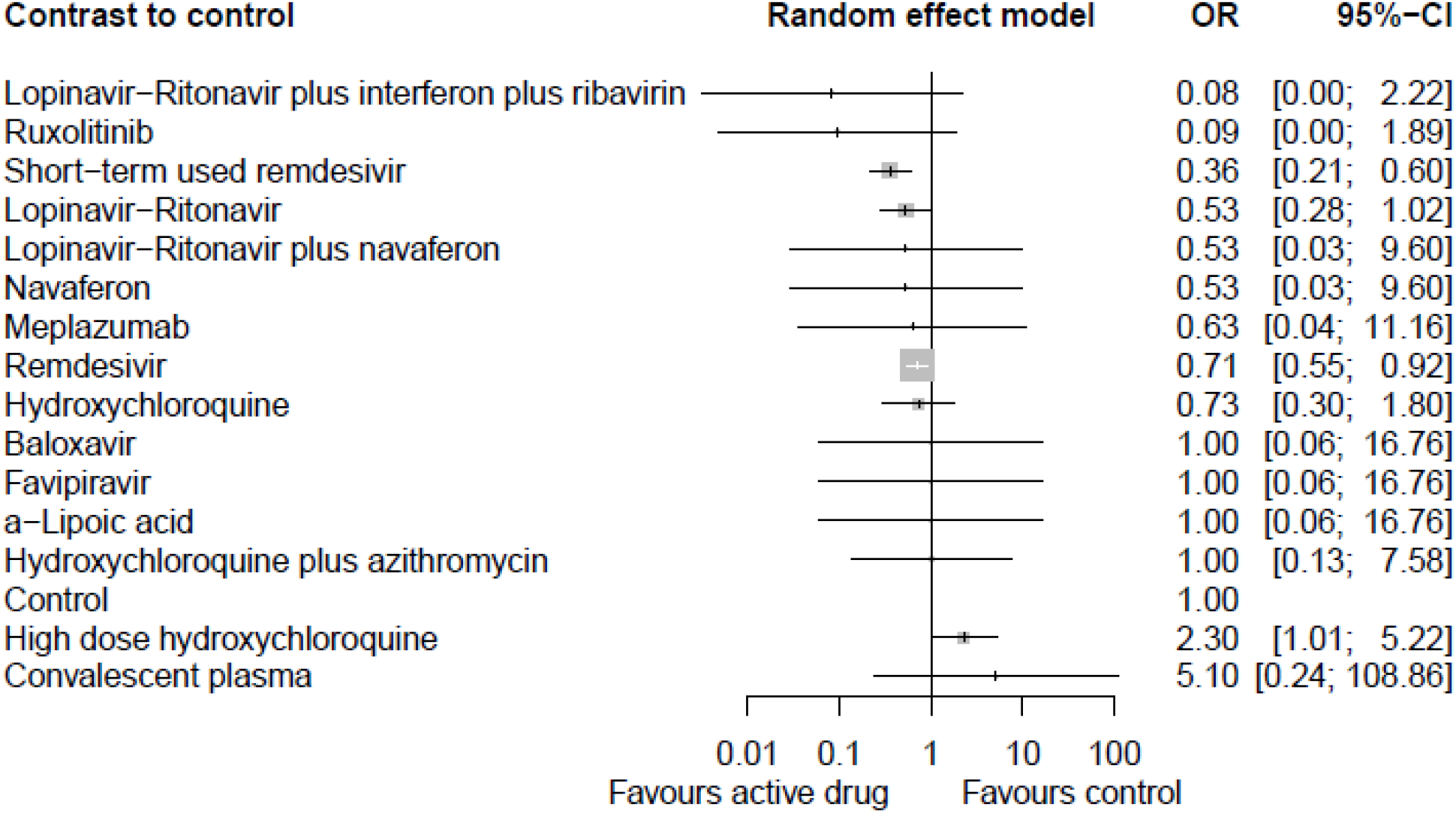
Network meta-analysis of safety of different pharmacological interventions. (A) Change in QTc interval (ΔQTc) from baseline (msec). (B) Proportion of patients experiencing QTc prolongation (>500ms or ΔQTc >60ms). (C) Fatal cardiac complication after hydroxychloroquine administration (torsades de pointes, cardiac arrest, and severe ventricular arrhythmia). (D) Non-cardiac serious adverse events. Effect estimates are presented in odds ratios (OR) and mean differences (MD) with 95% CI. Pharmacological agents are ranked by surface under the cumulative ranking curve (SUCRA) value. CAD = coronary artery disease. CHD = congestive heart disease. HQ = hydroxychloroquine.

### Fatal cardiac complications: Torsades de pointes, cardiac arrest, and severe ventricular arrhythmia

The associations between fatal cardiac complications and hydroxychloroquine, azithromycin, or hydroxychloroquine plus azithromycin therapy were analyzed (Figure 5C). Overall, treatment with hydroxychloroquine plus azithromycin showed a significant association (OR 2.25, 95% CI 1.27 to 3.99, low certainty) while others did not. We further subdivided the included studies based on prevalence of coronary artery disease (CAD) and congestive heart disease (CHD) at baseline. In studies in which >10% of the baseline population had CAD/CHD, the risk of fatal cardiac complication was statistically significantly higher in patients receiving hydroxychloroquine plus azithromycin. In studies in which <10% of the baseline population had CAD/CHD, no notable difference in incidence of fatal heart complication was observed in any treatment group.

### Non-cardiac serious adverse events

High dose (>600mg/day) hydroxychloroquine was associated with increased non-cardiac serious adverse events (Figure 5D). In contrast, there was a protective tendency with a decreased rate of adverse events with remdesivir compared to standard care (OR 0.71, 95% CI 0.55 to 0.92, high certainty).

### Subgroup and sensitivity analysis

The results of our subgroup and sensitivity analysis are reported in the Appendix pp 48-62. The assessments of other specific complications such as nausea/vomiting, diarrhea, hypoalbuminemia, anemia, leukopenia, lymphopenia, elevated AST/ALT, elevated CK, and increase total bilirubin are also presented in Appendix pp 52-54.

## Discussion

This is the first network meta-analysis (NMA) of pharmacological treatment for COVID-19. We comprehensively analyzed 31 active pharmacologic agents and their combinations in a large-scale analysis incorporating 20212 confounder-adjusted patients. Our study included unpublished data to integrate recent investigations and avoid selection and publication bias, as done in previous studies^4-7^. We did not limit our inclusions to RCTs and incorporated observational studies as we deemed that, in this analysis, the inclusion of real-world evidence from non-randomized studies has the potential to add validity to certain findings^13^, provide additional information regarding low-to-moderate incidence adverse events^14-16^, and improve the density of the network^14^. Many previous NMAs included observational studies with this rationale^14-16 32 33^, but inclusion of observational studies to an NMA requires careful integration to avoid biases from these observational studies pervading the meta-analysis^34^; as such, we exclusively included cohort studies that adjusted for confounders through methods such as propensity score matching (PSM), inverse probability treatment weighting (IPTW), and regression model adjustment or established similarity in the baseline characteristics of the groups being compared so that such adjustments are not necessary or irrelevant.

### Statement of principal findings

Our conclusions support the use of individualized treatment strategies based on clinical setting and severity. For moderate and severe patients hospitalized in non-ICU settings, tocilizumab and anakinra were shown to reduce risk of progression to severe pneumonia or ICU admission. Both of these selective anti-inflammatory agents also showed survival benefit compared to standard care. Remdesivir was the only antiviral agent shown to prevent progression of disease to severe pneumonia or transfer to ICU, but it did not alter mortality rate for non-ICU patients. For ICU-based critically ill patients, high dose IVIG and tocilizumab may reduce mortality while corticosteroid was associated with increased mortality. Convalescent plasma and hydroxychloroquine, topics of much debate, were not shown to reduce mortality rate or prevent progression to severe disease in our analysis; however, they demonstrated benefit in promoting viral clearance.

### Implications for clinicians and policymakers

Our analysis showed that hydroxychloroquine was significantly associated with reduced time to viral clearance (Figure 4C). Although this result was not supported by a single RCT on this subject^35^, this RCT should be interpreted with caution due to the median 16 days of delay from symptom onset to the treatment; our analysis indicated that the effects of hydroxychloroquine may fade after 14 days of delay from symptom onset to treatment initiation (Figure 4E). It should be noted that hydroxychloroquine was not shown to reduce mortality rate or progression to severe courses. As the level of evidence (GRADE assessment) varies in certainty for these results, further prospective large randomized trials with early initiation of treatment may be warranted to establish firm conclusions.

The potential cardiotoxicity of hydroxychloroquine and azithromycin is a widely shared concern in treating COVID-19 with these medications. According to our quantitative synthesis, incidence of QT prolongation was significantly higher in the patients who received hydroxychloroquine plus azithromycin compared to those who received standard care (Figure 5B). In addition, this combination of hydroxychloroquine and azithromycin was also associated with increased rate of fatal cardiac complications such as torsades de pointes, cardiac arrest, and severe ventricular arrhythmia in the cardiac-impaired population with a pooled incidence of 12.2%; in comparison, the pooled fatal cardiac complications rate in healthy populations with preserved cardiac function was about 0%. Therefore, the use of hydroxychloroquine/azithromycin should be limited to patients with healthy cardiac function, and monotherapy should be preferred to combination therapy for patients with poor cardiac function (Figure 5C). It should also be noted that non-cardiac adverse events were significantly more frequent in high dose (>600mg/day) hydroxychloroquine monotherapy compared to standard care (Figure 5D); nausea, vomiting, and diarrhea that required discontinuation of the treatment were more frequent with high dose hydroxychloroquine intake. Strict monitoring should be implemented in all patients receiving hydroxychloroquine with or without azithromycin to maintain a tolerable safety margin.

The results of our study also showed the efficacy of remdesivir in reducing the progression of COVID-19 to more severe pneumonia or admission to the ICU (Figure 3E-F). This result was supported by a high certainty of evidence (Table 2) and was replicated in the sensitivity analysis that included only RCTs. Interestingly, non-cardiac serious adverse events occurred significantly less in patients who received remdesivir compared to the control group (Figure 5D). This may be explained by the preventive effect of remdesivir against progression to severe diseases as numerous clinical trials reported possible consequences of severe disease such as septic shock and acute kidney injury as non-cardiac serious adverse events.

Tocilizumab (monoclonal IL-6 receptor antibody), anakinra (IL-1 receptor antagonist), and IVIG were associated with significantly reduced mortality in COVID-19 patients (Figure 3A-D). These three agents are known anti-inflammatory agents that have been conventionally used in hyperimmune or autoimmune conditions; tocilizumab and anakinra have been used for the management of severe rheumatoid arthritis^36 37^ and juvenile idiopathic arthritis^38-40^, and IVIG was used for management of Kawasaki’s disease^41 42^, inflammatory muscle diseases^43 44^, and sepsis^45^. As there is accumulating evidence for an hyper-immune response characterized by the release of pro-inflammatory cytokines in severe and deceased Covid-19 patients^46-50^, suppression of the inflammatory response and potential cytokine storm with immune-modulatory therapies was proposed as a potential therapeutic target; the results of this network meta-analysis support the efficacy of these treatments. Effectiveness of anti-inflammatory agents (tocilizumab, anakinra, IVIG) and ineffectiveness of antiviral agents, except for remdesivir, in hospitalized COVID-19 patients suggest that the management should focus more on the immune response rather than viral mechanism itself. Although numerous studies reported consistent results on beneficial effect of such agents, the certainty of evidence for these agents are either low or very low because conclusions on tocilizumab, anakinra, and IVIG to date are all based on observational studies. Randomized controlled trials on these anti-inflammatory agents are required to confirm these findings and increase the level of evidence.

Corticosteroids, on the other hand, were associated with significantly increased risk of morality in critically ill COVID-19 patients (Figure 3C). It could be argued that the frequent use of corticosteroids on patients with more severe conditions may have skewed the results against corticosteroid use; however, the studies included in our synthesis have adjusted for confounders for mortality including severity of disease, implicating that the observed unsafe effect of corticosteroids in critically ill COVID-19 should not be neglected. The detrimental effects of corticosteroid on mortality is in line with previous studies that showed the higher mortality rate with adjuvant steroid use in HIN1 influenza-infected critically ill patients in the ICU due to increased rates of superinfection and non-selective suppression of immune responses^51-53^.

### Limitations

Our study has several limitations. First, some of the results were derived from a single study (i.e., Anakinra) or studies with high risk of bias. To account for such weakness in evidence, we assessed the certainty of evidence for each outcome using the GRADE framework as summarized in Table 2. Second, for certain treatment agents, many articles have been published among which only one or few have been included in our analysis (e.g. convalescent plasma). This is because we prospectively collected studies adhere to predefined inclusion criteria, and studies that did not adequately account for confounding or those prone to significant bias were filtered out. The excluded studies are listed and described in the supplementary appendix pp 171-174 with reasons for exclusion. Third, we included observational studies and unpublished data. While such inclusions may introduce biases into the final analysis, we judged the benefits overweigh the risks for reasons we mentioned in methods. Furthermore, we attempted to minimize biases by exclusively including observational studies with confounder-adjustment and further conducted sensitivity analyses in which the same analysis was performed using only RCTs or only published studies (Appendix pp 55-58). Lastly, some of the results derived from this NMA lacks the support of pairwise meta-analysis. However, the methodological power of NMA is credible as empirical evidence supported that NMAs were 20% more likely to provide stronger evidence against the null hypothesis than conventional pairwise meta-analyses^54^. Accordingly, our NMA can offer meaningful implications for guiding management of COVID-19 until future studies build up stronger evidence.

## Conclusion

Anti-inflammatory agents (tocilizumab, anakinra, and IVIG) and remdesivir may safely and effectively improve clinical outcomes of COVID-19. Widely used hydroxychloroquine provides marginal clinical benefit in improving viral clearance rates whilst posing both cardiac and non-cardiac safety risks. Only 20% of current evidence on pharmacological management of COVID-19 is on moderate/high evidence certainty and can be considered in practice and policy; remaining 80% are of low or very low certainty and warrant further studies to establish firm conclusions.

## Data Availability

Not applicable. This study is a systematic review and thus involve data available in open sources and publications.

## Contributors

MS Kim and MH An contributed to the study concept and design. MS Kim, MH An, and WJ Kim identified and acquired relevant trials and extracted data. MS Kim and WJ Kim drafted the protocol for this study. MS Kim and MH An analyzed and interpreted the data. MS Kim and MH An wrote the first draft of the manuscript. T Hwang contributed to the interpretation of data and critical revision of the manuscript. All authors saw and approved the final submitted version.

## Ethical approval

Not required

## Data sharing

Not applicable

## Dissemination declare

Not applicable

## Transparency

The manuscript’s guarantor (MSK) affirms that this manuscript is an honest, accurate, and transparent account of study being reported; that no important aspects of the study have been omitted; and that any discrepancies from the study as planned have been explained.

